# A new approach to estimating HIV incidence and the size of the infected and undiagnosed population in high immigration settings

**DOI:** 10.1101/2025.02.14.25322154

**Authors:** Amber Kunkel, Françoise Cazein, Florence Lot, Julie Muzzolon, Nikos Pantazis, Amadou Alioum

**Affiliations:** Unité VIH-hépatites B/C-IST, Direction des maladies infectieuses, Santé publique France, Saint-Maurice, France; Master Statistique, Modélisation et Science des Données, Université Claude Bernard Lyon 1, Lyon, France; Dept. of Hygiene, Epidemiology and Medical Statistics, School of Medicine, National and Kapodistrian University of Athens, Greece; Univ. Bordeaux, National Institute for Health and Medical Research (INSERM) UMR 1219, Bordeaux Population Health, Bordeaux, France

**Author notes:** Corresponding author: Amber Kunkel, 12 Rue du Val d’Osne, All. Vacassy, 94410 Saint-Maurice, France.

## Abstract

Most current approaches to estimating HIV incidence and the size of the undiagnosed population do not account for pre-migration infections, although these represent a substantial fraction of new diagnoses in some European countries. We propose a new approach to estimating these indicators that explicitly accounts for migration. First, we use an existing Bayesian model to estimate delays between infection and diagnosis for each newly diagnosed individual, and classify those born abroad into pre- or post-migration infections. Second, for native-born individuals and those with post-migration infections, we use a repurposed regression model to describe the global distribution of delays between infection and diagnosis, and estimate HIV incidence. For people with pre-migration infections, the same model is used to describe the delays between migration and diagnosis, and subsequently estimate the number of arriving migrants with undiagnosed HIV. Finally, these estimates are combined to estimate the number of people in the country with undiagnosed HIV. When applied to simulated data inspired by the French HIV surveillance system, the new model appropriately corrects for pre-migration HIV infections and produces estimates of HIV incidence, arrival of people with undiagnosed HIV, and the size of the undiagnosed population that are similar to the true simulated values.

## Introduction

Estimating HIV incidence and the number of people with undiagnosed HIV infection is necessary to evaluate countries’ progress towards the United Nations Sustainable Development Goal to end AIDS by 2030 ^1^. These estimations are not trivial, owing to the long delay between HIV infection and development of symptoms and the complex dynamics of HIV testing behavior. Several mathematical models have been proposed to address these challenges, including back-calculation approaches ^2–4^ and models based on recent infection assays ^5, 6^.

Immigration of people with pre-existing undiagnosed HIV infection presents another challenge to HIV indicator estimation and interpretation. Pre-migration infections can account for a substantial proportion of new HIV diagnoses in some settings. In France, for example, 56% of people newly diagnosed with HIV in 2022 were born abroad, among whom estimated rates of pre-migration infections range from approximately 40% for men who have sex with men (MSM) to 50-65% for adults born in sub-Saharan Africa ^7–9^. The majority of existing HIV incidence estimation methods do not account for pre-migration infections. Back-calculation methods technically estimate HIV incidence for all individuals eventually diagnosed within a country, regardless of whether they were infected prior to migration or not. Methods based on recent infection testing may more closely approach post-migration HIV incidence, but typically cannot distinguish between pre- and post-migration diagnosis delays.

Unlike most other incidence estimation approaches, the ECDC HIV Modeling Tool is being adapted to exclude people with likely pre-migration infection from HIV incidence estimates ^10^. The new optional migration module of the ECDC HIV Modeling Tool uses a pre-existing Bayesian model to identify and exclude people with likely pre-migration infection from its incidence estimates. The same model was previously used to estimate the proportion of people with pre-migration infection in the aMASE study and in European surveillance data of new HIV diagnoses ^11–13^. The model works by estimating a posterior distribution of possible infection dates for each person newly diagnosed with HIV, based on information such as their CD4 count, HIV-RNA viral load, date of last negative test, demographic/clinical characteristics and presence of AIDS symptoms at the time of diagnosis. These estimated infection dates are then compared with the person’s date of migration to classify the infection as occurring pre- or post-migration.

In this paper, we propose to go a step further and use these estimated distributions of infection dates directly to estimate other key indicators, including the number of people living with undiagnosed HIV. Because we can identify newly diagnosed people with an estimated pre-migration infection and know their date of immigration, the proposed model also allows us to estimate the number of people immigrating each year with undiagnosed HIV infection. The sole data requirement is a surveillance dataset of new HIV diagnoses that includes several consecutive years of individual-level data on date of HIV diagnosis, key demographic variables, CD4 cell count, clinical stage, date of last negative test, and date of migration. The performance of this new model is evaluated using simulated data. Although the primary advantage of this proposed approach is its explicit accounting for migration, and the subsequent improvement in HIV indicator estimates when a substantial fraction of new HIV diagnoses acquired their infections abroad, it also performs well on simulated data with no immigration.

## Methods

This section first provides an overview of the main steps of the estimation procedures in our proposed approach and then details each step.

### Brief overview of the proposed approach

We start with a dataset containing all new HIV diagnoses (N_diag_) in the country of interest in the period from years Y_min_ to Y_max_. The time between Y_min_ and Y_max_ should be sufficiently long so that the majority of people infected in Y_min_ will have been diagnosed by Y_max_; we suggest considering a period of at least 10 years. This dataset must include the following variables measured at the time of diagnosis: diagnosis date, age, sex, probable mode of HIV acquisition, place of birth (continent), CD4 cell count, presence or absence of AIDS, date of last negative HIV test (if any), and immigration date (only for migrants). We assume this dataset contains no missing data after performing a prior optional multiple imputation procedure with n_imp_ imputations. The total size of the dataset prior to the estimation of infection times is thus N_diag_*n_imp_, which can be seen as a concatenation of n_imp_ pseudo-complete data sets of N_diag_ individuals each. If there are no missing values in the aforementioned variables, the imputation procedure is not required thus the total size of the dataset is N_diag_.

A posterior distribution of plausible infection times for each individual in this dataset is derived using a modified version of the model described in ^12^, and n_draw_ draws of infection times from his or her posterior distribution are taken using rejection sampling. The final size of the data is then n_draw_*n_imp_*N_Diag_. For the following steps, each of the n_draw_*n_imp_ instances of the original dataset of size N_diag_ is treated independently, as if there were n_draw_*n_imp_ imputations and will be referred to as an imputed dataset. It is important to stress again here that if there are no data missing in the initial data of N_diag_ individuals, and that there is no need to make multiple imputations in the classical sense, then we consider that we have n_draw_ datasets that we will treat as “imputed” data, as described in ^12^.

For each of the n_draw_*n_imp_ imputed datasets of size N_diag_, we estimate the delay distribution between infection and diagnosis and subsequently HIV incidence using methods originally proposed for handling AIDS reporting delays ^14^. To exclude pre-migration infections from the incidence estimate, only those individuals for whom the estimated date of infection (in the specific imputed dataset) occurred after the date of immigration are included in this calculation. In parallel, we estimate the delay distributions between immigration and diagnosis for individuals with an estimated date of infection prior to the date of immigration and, in place of incidence, the number of people immigrating each year with undiagnosed pre-migration HIV infections.

Next, for each imputed dataset, we estimate the size of the undiagnosed population as follows. For people infected in the country of interest, we start with the total number of individuals infected fromY_min_ to Y_max_ and subtract those already diagnosed by the end of Y_max_. To include infections prior to Y_min_, we project forward the number of people diagnosed each year that were infected prior to Y_min_ assuming a log-linear trend. This approach is similar to that described in ^15^. A similar approach is used for people with pre-migration infections, with reference to the date of immigration rather than the date of infection.

Finally, the estimated incidence and the size of the undiagnosed population are combined across all imputations and draws from the posterior distribution using Rubin’s rules, as in ^12^.

### Dates of infection

A Bayesian model integrating subject-specific information on CD4, viral load, clinical stage, and previous negative HIV tests along with estimates from a model fitted to data from individuals with well estimated dates of HIV seroconversion, was previously developed to estimate the proportion of infections occurring pre- and post-migration in the aMASE study ^12, 13^, and subsequently applied to European surveillance data ^11^. A key feature of this model is the generation of a distribution of estimated infection times for each individual. Within the model, a uniform prior distribution of estimated time from infection to diagnosis is defined with a minimum delay of >0 years (we used 0.01 year) and a maximum determined by the date of the last negative HIV test, the age (assuming infections occur in individuals ≥10 years old in the original model, or ≥15 years in our version), and the calendar year (assuming all infections occur after 1980). A likelihood function is defined according to the individual’s CD4 cell count, viral load, and AIDS status at the time of diagnosis.

We have made a few modifications to the original model (Appendix A.1). First, to account for the fact that a single individual cannot have more than one first-time HIV diagnosis, we multiply the original likelihood function by a survival function S_diag_(t) representing the probability of not yet being diagnosed by time t. This survival function is approximated using a method previously described in ^5, 6^. In this method, the AIDS incubation period is assumed to follow a gamma distribution and the time from HIV infection to diagnosis (prior to AIDS) is assumed to follow an exponential distribution with mean 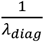. The parameter *λ*_*diag*_ is then estimated from the proportion of HIV diagnoses at the AIDS stage (*p*_*aids*_). Similar to ^5^, the AIDS incubation period for all individuals in the dataset is approximated using a Gamma(shape=5.251, scale=1.974) distribution. Separate estimates of *λ*_*diag*_ are made in strata defined by probable mode of HIV acquisition and place of birth. Specifically, within each covariate stratum *k*,

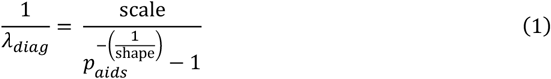

The survival function then has the form S_diag_(t)=exp(−*λ*_*diag*_t). As in ^5^, we apply this approach only to first-time testers, and assume that testing dynamics differ for individuals with a prior negative test.

Second, to improve the speed of the model estimating infection dates, we use only CD4 count, not viral load. We also group similar individuals by rounding CD4 count to the nearest multiple of 50 (max 1500), age to the nearest multiple of 5 years, calendar year to the nearest multiple of 5 years, and the maximum length of the prior distribution to the nearest quarter (if <5 years) or year (if ≥5 years). These changes may not be necessary in some settings, but improve the model’s runtime on large datasets.

As in ^12^, n_draw_ estimated infection times are drawn from the posterior distribution for each (imputed) individual in the original database using rejection sampling. The full dataset is thus augmented to n_draw_ times its original number of rows. In the following steps, each subset of size N_diag_ consisting of a single imputation and estimated infection time per individual is treated independently. At this stage, we are able to separate individuals in each imputed dataset into those infected before migration (pre-migration infections) or those infected after migration (post-migration infections), depending on whether their estimated date of infection is before or after their observed (or imputed) date of arrival (if relevant).

### HIV incidence

To estimate HIV incidence in the country of interest, we first exclude all individuals with an estimated infection time prior to the date of migration (if any). These are considered pre-migration infections. The remaining data are grouped according to the delay in quarters between infection and diagnosis, truncated at a pre-defined value *m* less than the total number of quarters between Y_min_ and Y_max_, and any covariates to be included in the modeled delays between infection and diagnosis, such as sex, probable mode of HIV acquisition transmission, or year of infection.

The methods used to estimate HIV incidence were previously developed to estimate distributions of AIDS reporting delays and to correct AIDS incidence for reporting delays ^14^. The first step is to estimate the diagnosis delay distribution F(t).

Within covariate strata k, Y_jk_ is defined as the number of cases with diagnosis delay = t_j_ (0 ≤ j ≤ m) and n_jk_ is defined as the number of cases with diagnosis delays ≤ t_j_ and truncation times (i.e. maximum observable delays) ≥ t_j_. The conditional probability p_jk_ is the probability of a diagnosis delay = t_j_, given the delay is ≤ t_j_. Then Y_jk_ is assumed to follow a binomial distribution Y_jk_ ~ Binomial(n_jk_, p_jk_). The binomial probabilities for j≥1 are modeled using a generalized linear model with a complementary log-log link:

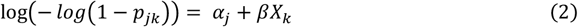

The terms α_j_ allow the probabilities to differ for each possible delay (in quarters) and the vector of regression coefficients *β* allows for the inclusion of covariates *X*_*k*_, which may include the quarter or year of infection and any stratifying variables such as probable mode of HIV acquisition or place of birth. This model can be easily fitted using software for GLMs. The cumulative probability that the diagnosis delay is less than or equal to *t*_*s*_ for strata k with covariates *X*_*k*_ is then ^14^:

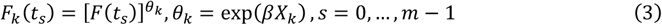

where 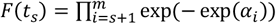 is the diagnosis delay distribution when all of the covariates are 0.

We propose a method to estimate the variance *var* 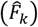 of the estimated diagnosis delay distribution 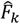 using the delta method (Appendix A.2).

Based on the estimated diagnosis delay distribution, we next derive the estimated HIV incidence among people with covariate *X*_*k*_ for each quarter *q* from Y_min_ to Y_max_. To do so, we divide the number Z_k_(q) of people diagnosed through the end of Y_max_ with an estimated infection date in *q* by the probability an infection occurring in *q* would be diagnosed before the end of Y_max_ with last quarter q_max_:

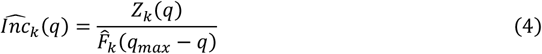

Following ^14^, the variance of the incidence in each quarter is approximated using the delta method :

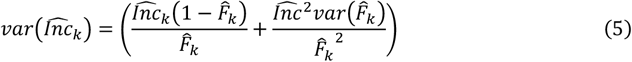

Details of the calculation of *var*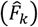 using the delta method are given in Appendix A.2.

Finally, the incidence per year per covariate stratum k is calculated by adding the incidence for each quarter in that year, and the variance is approximated by adding the variance for each quarter within the year. The overall incidence per year and its variance are similarly calculated as the sum of all strata.

Once annual HIV incidence and its variance are estimated for each imputation and infection time draw, overall point estimates and confidence intervals of annual HIV incidence are derived according to Rubin’s rules.

### Migration of individuals with undiagnosed HIV infection

The method used to estimate the number of migrants with undiagnosed HIV infection arriving in the country is very similar to the method used to estimate HIV incidence. For this estimation, the data is restricted to only those rows consisting of new HIV diagnoses infected prior to immigration (pre-migration infections). In this case, it is the distribution of time between immigration (instead of infection) and HIV diagnosis that is estimated. Delays are expressed in years rather than quarters, as surveillance data often includes only year of immigration. The distributions *G*_*k*_are estimated to depend on covariates *X*_*k*_ defining strata k, as in the case of the estimates of the distributions *F*_*k*_ of delays between infection and diagnosis presented above.

The number of arriving migrants with undiagnosed HIV infection for each year y from Y_min_ to Y_max_ for each stratum k is estimated by the number W_k_(y) of people diagnosed through the end of Y_max_ with an immigration year of *y* divided by the probability an undiagnosed infection in a migrant arriving in *y* would be diagnosed before the end of Ymax:

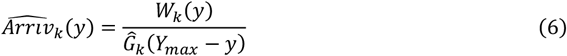

The variance is estimated using the same procedures as for HIV incidence.

### Size of the population living with undiagnosed HIV

To estimate the number of people living with undiagnosed HIV at the end of Y_max_, we assume that native-born individuals and those infected post-migration contribute undiagnosed person-time from the date of infection to the date of diagnosis, and that individuals with pre-migration infections contribute undiagnosed person-time from the date of immigration to the date of diagnosis. These estimates can be made separately within covariate strata k describing patient characteristics like country of birth or probable mode of HIV acquisition, but no longer including year of infection. We describe below the procedures for estimating the number of undiagnosed people living with HIV infected in the country of interest versus prior to migration. These two estimates are added together to produce the total size of the undiagnosed HIV epidemic.

### Size of the population living with undiagnosed HIV: people infected within the country

For individuals infected within the country of interest, the number of people infected at each quarter *q* from Y_min_ to Y_max_ and still undiagnosed at the end of Y_max_ is estimated as:

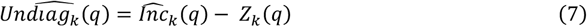

Quantities are defined as in equation (4), except that infection times are not truncated after *m* quarters in the estimation of *Z*_*k*_*(q)*. As *Z*_*k*_*(q)* is treated as fixed within each imputation, the within-imputation variance of the number of undiagnosed individuals infected in each quarter *q* is assumed to equal the variance of the estimated incidence for that quarter. Finally, the overall number of people infected from Y_min_ to Y_max_ and undiagnosed by the end of Y_max_ is calculated by adding the number of individuals infected in each quarter remaining undiagnosed, and the variance is approximated by adding the variance for each quarter. Estimates are combined across imputations using Rubin’s rules.

To calculate the number of individuals infected prior to Y_min_ and still undiagnosed at the end of Y_max_ for strata k with covariates X_k_, we used an approach similar to ^15^. We observed in both simulated data and surveillance data from France that the number of people infected prior to Y_min_ and diagnosed each year t from Y_min_ to Y_max_, M(t), followed an approximately log-linear trend (Appendix A.3). We thus fit a Poisson model:

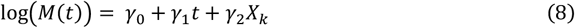

Using this model, the number of people infected prior to Y_min_ and diagnosed each year t is projected forward to t=Y_min_+30, after which we assume no further diagnoses. The sum of estimated diagnoses occurring from Y_max_+1 to Y_min_+30 is taken to be the number of undiagnosed individuals infected prior to Y_min_ remaining undiagnosed at the end of Y_max_. Its within-imputation variance is estimated by taking 100 bootstrap samples of the parameters of the Poisson model. To improve stability of estimates for small strata, we further assume that the number of people infected prior to Y_min_ and diagnosed after Y_max_ could not exceed three times the number of people infected prior to Y_min_ and diagnosed between Y_min_ and Y_max_. This assumption should be conservative if at least 10 years of data are used, given the median time to AIDS diagnosis is approximately 8-11 years ^16^. Estimates are again combined across imputations using Rubin’s rules.

The number of undiagnosed individuals infected in the country is then calculated as the sum of the number of individuals infected prior to Y_min_ and still undiagnosed at the end of Y_max_ and the number of individuals infected between Y_min_ and Y_max_ and still undiagnosed. The variance is approximated as the sum of the variance of these two estimates.

### Size of the population living with undiagnosed HIV: people with pre-migration infections

Similar procedures are used to estimate number of undiagnosed individuals infected prior to migration. In this case, the number of undiagnosed arrivals calculated in (6) is substituted for HIV incidence in equation (7) above, which is applied to people arriving in the country each year between Y_min_ and Y_max_.

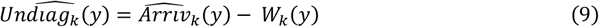

An approximate log-linear trend in the number of people arriving before Y_min_ with pre-migration infections and diagnosed between Y_min_ and Y_max_ was again observed (Appendix A.3). The number of people with pre-migration infections who arrived prior to Y_min_ and remained undiagnosed by the end of Y_max_ was estimated by projecting this trend forward to Y_min_+30 using a Poisson regression model similar to equation (8), after which we assume no further diagnoses. The sum of estimated diagnoses occurring from Y_max_+1 to Y_min_+30 is taken to be the number of undiagnosed individuals arrived prior to Y_min_ remaining undiagnosed at the end of Y_max_.

## Simulation study

The methods described above were implemented in R version 4.3.1. These codes, as well as the simulated data described below, are available as supplementary material.

### Construction of a simulated dataset

To test the model, we created a simulated dataset of new HIV diagnoses in a given European country inspired by the HIVSynthesis model ^17–20^ and other simulated data of new HIV diagnoses ^3^. The parameters of this dataset were based on observed data from the HIV surveillance system in France, and its construction is described in detail in Appendix A.4.

Briefly, the number of HIV infections per year is assumed known from its presumed start in 1982 through 2022. The shape of this curve is inspired by previous estimates of HIV incidence in France ^2^. To simplify the simulation, all simulated individuals are assumed to be MSM born in Europe, with a pre-specified age distribution. We assume 50% of individuals are born outside of the country. For comparison, in 2022, 56% of new HIV diagnoses in France were born abroad ^7^.

Following the HIVSynthesis model ^17^, we simulated individual viral load and CD4 trajectories over time (in quarters) for each new infection. The development of symptoms and AIDS are assumed to depend stochastically on CD4, viral load, and age.

Among first-time testers (i.e. those without a previous negative HIV test at the time of HIV diagnosis), we assume the date of diagnosis is determined according to probabilities that vary according to clinical stage. The annual rate of diagnosis in asymptomatic individuals varies by calendar year. Starting from 1995, a higher probability is assigned to individuals in the first three months of HIV infection, who may have symptoms of primary HIV infection or knowledge of a high-risk exposure. A higher probability is also assigned to individuals with symptoms, and particularly those with AIDS-defining conditions. Among individuals with a previous negative test, we assumed HIV testing follows a Poisson process, similar to ^3^. In contrast to first-time testers, a date of diagnosis is thus determined for each individual prior to simulating his trajectories of viral load, CD4 count, and symptoms. For both first-time and repeat testers, we allow a temporary drop in the probability of diagnosis (by 30-50%) during the years 2020-2021 to reflect possible disruptions caused by the COVID-19 pandemic. We also created a second version of the simulated dataset that does not include this COVID-associated drop in diagnoses.

Dates of immigration were simulated for 50% of individuals in the dataset assumed to be born abroad. We aimed for the distribution of the time between immigration and HIV diagnosis in the simulated data to be similar to that observed for MSM born in other European countries in the French surveillance data.

The full simulated dataset included 338,652 rows, each representing a single person assumed infected with HIV from 1982-2022 and diagnosed in the country. For simplicity, we tested the model on a smaller dataset created by taking a random sample of 50,000 of these rows, approximately 15% of the original dataset. We assumed the dataset was complete with no missing data, so no imputations were applied. For the figures presented here, we drew n_draw_=15 samples from the infection times distribution of each simulated individual.

### Comparison with ECDC HIV Modeling Tool

One of the main models currently used to estimate HIV incidence by countries in Europe is the ECDC HIV Modeling Tool ^10^. This approach, which has a user-friendly online interface, is based on a multi-state back-calculation model of CD4 decline over time, and models HIV incidence using cubic splines ^4^. We exported the simulated datasets to use with this model, assuming data availability matching that of the HIV surveillance system in France: new HIV and joint HIV/AIDS diagnoses per year starting in 2003, with CD4 counts available from 2008. Although this tool has recently introduced an option of removing pre-migration infections from incidence calculations, we did not test this functionality, as it was not yet fully operational at the time of these analyses. Instead, we compared the performance of the ECDC HIV Modeling Tool and our new model on a dataset with no migration.

The ECDC HIV Modeling Tool allows one to define a functional form for the change in diagnosis probabilities over time. We originally wished to assume a linear trend in diagnosis probabilities for 2012-2022, with a temporary drop in 2020. However, the ECDC tool does not permit this sort of temporary discontinuity. Instead, we tested three other assumed trends in diagnosis probabilities: first with a linear trend from 2012-2020 and a jump to a new constant value in 2020, second with a linear trend in 2012-2022, and third with a linear trend from 2012-2020 and a jump to a new linear trend in 2020. We also applied these assumptions to our new model, with a key difference: while the structure of the ECDC model allows a direct change in the diagnosis probabilities for a given year, the new model describes rather the distribution of diagnosis delays for people infected in that year.

### Model performance on simulated data

On a simulated dataset with no immigration, the new model’s performance is largely comparable to that of the ECDC HIV Modeling Tool, with a few exceptions that generally favor the new model (Table 1, Figure 1). The true simulated size of the undiagnosed population in 2022 is within the confidence intervals of both models, excluding one scenario for the ECDC model (Test 1, assuming a linear trend in diagnosis probabilities from 2012-2020 and a jump to a new constant value in 2020) (Table 1). Both models reconstruct reasonably well the decline in simulated incidence in the beginning of the period from 2012 to approximately 2019, though the true simulated values more frequently lie within the confidence bands of the new model than of the ECDC model (Figure 1). The stabilization in incidence from 2019-2021 is more visible with the new model. The new model estimates for the period 2012-2021 are highly similar across all scenarios. In contrast, the ECDC HIV modeling tool appears more sensitive to the assumed trends in diagnosis probabilities, particularly when applied to the dataset with a temporary drop in diagnoses in 2020-21. Both models struggle somewhat to estimate the incidence in 2022 for that dataset. Compared to a true simulated value of 731, the ECDC model estimates range from 732 to 947, with two of the three confidence intervals containing the true value. The new model estimates range from 799 to 888 and, although all of the confidence intervals contain the true value, the width of these intervals is wide.

**Table 1:**
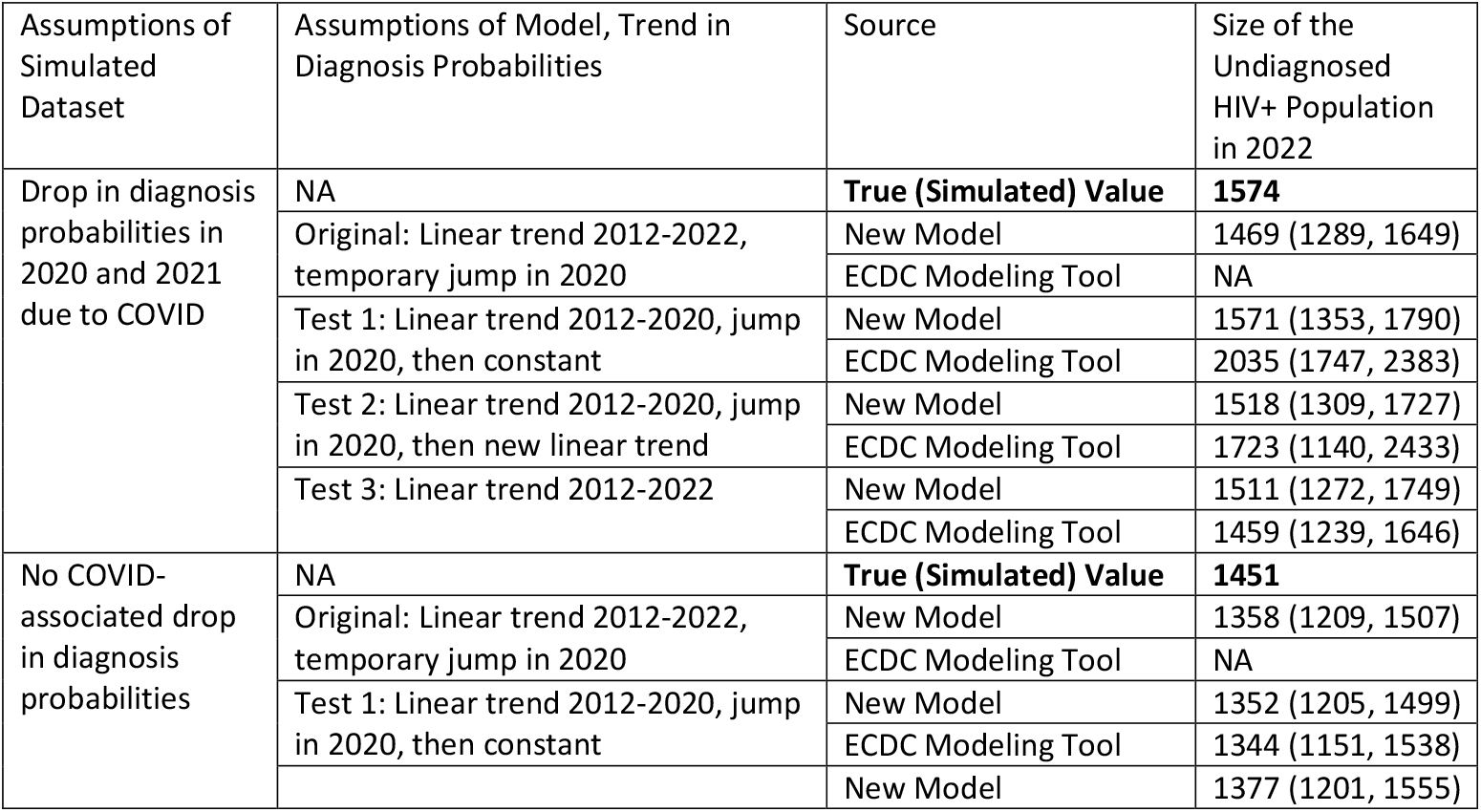
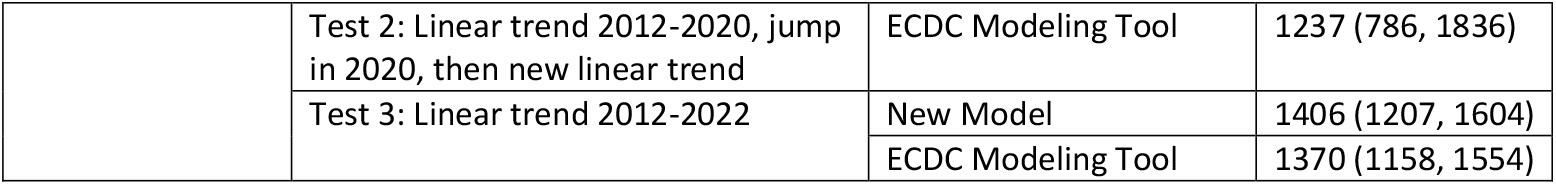
Comparison of the new model and the ECDC HIV modeling tool with four different diagnosis probabilities trend scenarios on a simulated dataset with no immigration – size of the undiagnosed HIV+ population in 2022.

**Figure 1.**
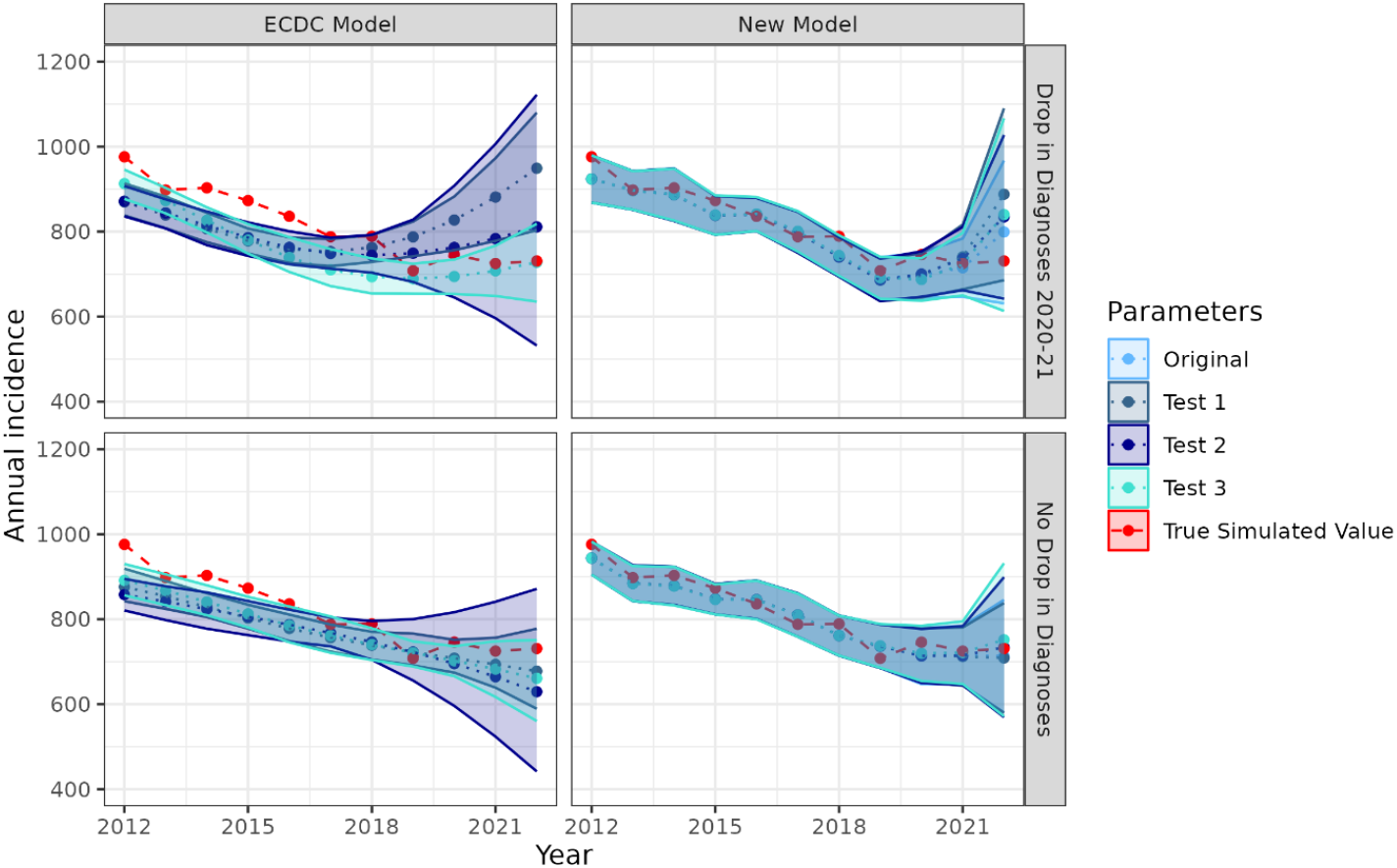
Comparison of the new model and the ECDC HIV modeling tool with four different diagnosis probability trend scenarios on a simulated dataset with no immigration, with and without an assumed drop in diagnoses during the COVID-19 pandemic – HIV incidence, 2012-2022. Original scenario: linear trend in diagnosis probabilities in 2012-2022, temporary jump in 2020. Test 1: linear trend in diagnosis probabilities in 2012-2020, jump in 2020, then constant. Test 2: linear trend in diagnosis probabilities in 2012-2020, jump in 2020, then new linear trend. Test 3: linear trend in diagnosis probabilities in 2012-2022

When applied to a simulated dataset that includes immigration of people with pre-migration HIV infections, the new model continues to produce largely acceptable estimates of HIV incidence and the size of the undiagnosed HIV population (Table 2, Figure 2). It is also produces reasonable estimates of the number of immigrants arriving with undiagnosed HIV each year from 2012-2022 (Table 2, Figure 3). A simplified version of the model that ignores migration (i.e. assumes all HIV infections occur post-migration), however, over-estimates both HIV incidence and the size of the undiagnosed HIV population in the country of interest (Table 2, Figure 2).

**Table 2:** Model performance on a simulated dataset including pre-migration infections – HIV incidence, number of immigrants with undiagnosed HIV, and size of the undiagnosed HIV+ population in 2022.

| Assumptions of Simulated Dataset | Source | HIV incidence in 2022 | Immigrants arriving with undiagnosed HIV in 2022 | Size of the undiagnosed HIV+ population in 2022 |
| --- | --- | --- | --- | --- |
| Drop in diagnosis probabilities in 2020 and 2021 due to COVID | **True (Simulated)** | **625** | **121** | **1178** |
|  | New Model Estimate | 711 (559, 863) | 103 (84, 122) | 1230 (1068, 1392) |
|  | New Model Estimate, Ignoring Migration | 799 (631, 967) | 0 | 1469 (1289, 1649) |
| No COVID-associated drop in diagnosis probabilities | **True (Simulated) Value** | **637** | **136** | **1135** |
|  | New Model Estimate | 622 (496, 748) | 117 (100, 134) | 1118 (979, 1259) |
|  | New Model Estimate, Ignoring Migration | 713 (581, 845) | 0 | 1358 (1209, 1507) |

**Figure 2.**
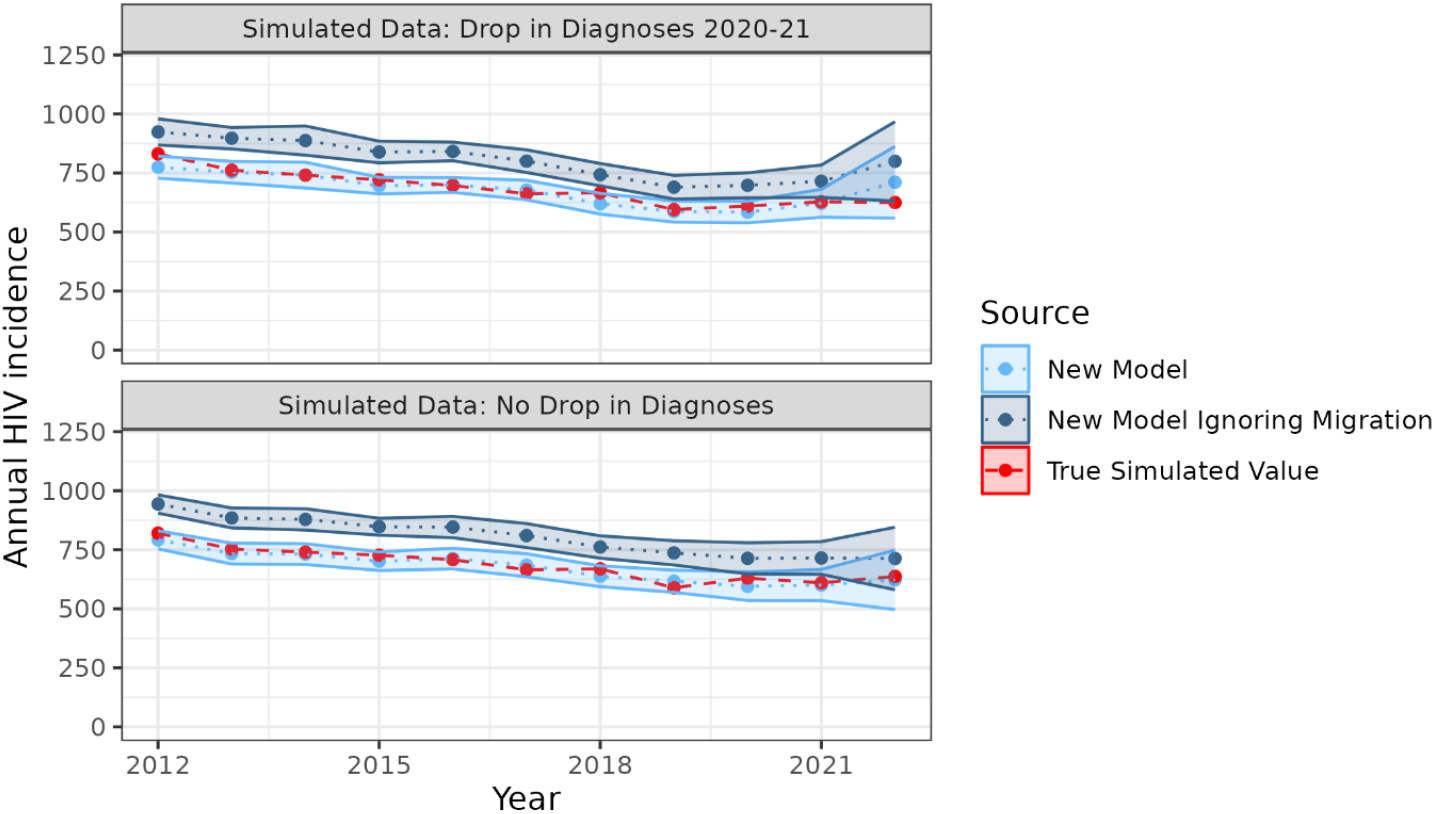
Model performance on a simulated dataset including pre-migration infections – HIV incidence, 2012-2022

**Figure 3.**
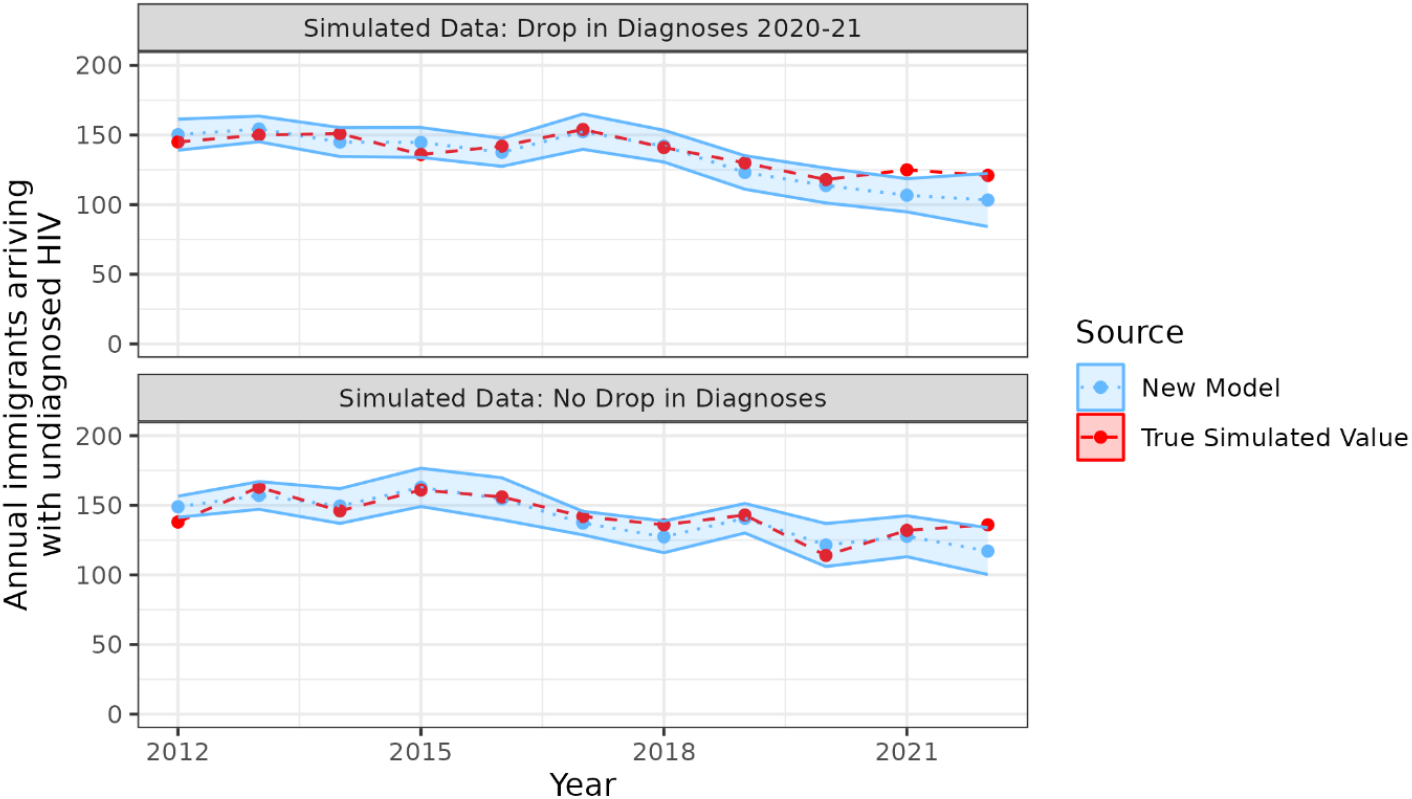
Model performance on a simulated data including pre-migration infections – immigration of people with undiagnosed HIV, 2012-2022

## Discussion

We have presented here a new method for estimating HIV incidence and the number of people with undiagnosed HIV infection that combines two models previously developed for other purposes: first, a Bayesian model that allows for the estimation of individual-level infection time distributions, and second, a model to estimate delay distributions and thereby to correct incidence for reporting delays. The primary advantage of this new method is that it explicitly accounts for migration of people with undiagnosed, pre-existing HIV infections, without requiring any data external to the routine HIV surveillance data (e.g. migratory flows, estimates of HIV prevalence in the country of origin). In addition, it also demonstrates good performance on simulated data with no migration compared to another commonly used method, the ECDC HIV Modeling Tool.

The underlying approach taken in this paper is conceptually similar to that used by Song et al. to estimate HIV incidence and the size of the undiagnosed HIV+ population in the United States ^15^. First, dates of infection are estimated for each notified individual; second, methods used for AIDS reporting delays are adapted to estimate HIV incidence; and finally, an approach based on regression and forward projection is used to account for undiagnosed HIV infections infected prior to the period for which incidence is estimated. However, the details of the methods used at each step differ significantly between the two models. For example, the Bayesian model used here to estimate delays between infection and diagnosis accounts for previous negative tests, CD4 cell count, and AIDS status at diagnosis, compared to the simpler CD4 depletion model used in Song et al. Most importantly, the approach by Song et al. does not account for pre-migration infections.

The motivation for the development of this method was to improve HIV indicator estimation in France, where 56% of new HIV diagnoses in 2022 were born abroad, and of these 40-65% likely acquired their infections prior to migration ^7–9^. The performance of the model on simulated data here shows that not accounting for migration can lead to substantial over-estimates in both HIV incidence and the number of people with undiagnosed HIV infections. A key variable needed to account for pre-migration infections is the date of immigration, which in France has only been collected since 2012. When tested on simulated data, the new model here showed good performance on a dataset restricted from 2012-2022. An additional advantage of this approach is that it can be easily integrated into existing data correction and analysis procedures such as those used in France, particularly multiple imputation to account for missing data.

The model presented here nevertheless has several limitations. First, we assume that the parameters describing HIV progression in the model used to estimate dates of HIV infection (based on the CASCADE consortium) can also be applied to new HIV diagnoses in the population of interest, conditional on key covariates like continent of birth, age, and sex. Second, although the model for estimating delay distributions allows these distributions to vary according to year of infection, it may not fully account for sudden changes occurring in a given year of diagnosis, as occurred during the COVID-19 pandemic. Third, incidence estimates result in relatively wide confidence intervals for the most recent year (2022 in our simulation), what also happens with all back-calculation approaches. These intervals are particularly wide as we did not assume any functional form or apply any smoothing procedures to the estimated incidence over time. As seen in the performance of the ECDC HIV Modeling Tool here on simulated data, such smoothing functions can lead to smaller confidence intervals but also have more unexpected effects on the shape of the estimated incidence curve over time. Fourth, this remains a relatively simple approach to accounting for migration in that it assumes migration occurs at a single point in time, ignoring the fact that people may go back and forth between countries over time, and ignoring any migration out of the country of interest.

In conclusion, we propose a new method for estimating HIV epidemic indicators in that accounts for pre-migration infections by excluding them from the calculation of HIV incidence and including them in estimates of size of the undiagnosed population only from the time of their migration. This approach can prevent over-estimation of HIV incidence and the size of the undiagnosed population that can occur when ignoring place of HIV acquisition. These estimates can be applied using HIV surveillance data on new HIV diagnoses, as long as certain key variables (year of migration, date of diagnosis, CD4 count, previous negative testing, AIDS status, region of birth, age, sex, probable mode of HIV acquisition) are available over a sufficient period of time (2012-2022 for the analyses presented here). Routine yearly application of these methods is planned in France. An example simulated dataset and R code are provided as supplementary material to assist other countries that may be interested in implementing a similar approach.

## Supporting information

Supplemental Data and Programmes

## Data Availability

The simulated data presented here and programs needed to run the model are included as supplementary material.

