## Supplemental Appendices for "A new approach to estimating HIV incidence and the size of the infected and undiagnosed population in high immigration settings"

A.1 Distribution of times between infection and diagnosis

A.2 Derivation of a variance formula for the delay distributions between infection or migration and diagnosis

A.3 Approximately log-linear decline in diagnoses of people infected/immigrated prior to $Y_{min}$

A.4 Detailed description of simulated dataset

### A.1 Distribution of times between infection and diagnosis

To account for the concurrent risk of an HIV diagnosis before AIDS, we have modified the likelihood proposed in (1) to include the probability of not being HIV diagnosed by a given time.

For people diagnosed prior to the AIDS stage of HIV infection, the distribution of times between infection and diagnosis are described as follows:

$$f(w_i|y_i^c, T_i > w_i) = \frac{f(y_i^c|w_i)S_{aids}(w_i)S_{diag}(w_i)f(w_i)}{\int_0^{u_i} f(y_i^c|w_i)S_{aids}(w_i)S_{diag}(w_i)f(w_i)dw_i}, 0 < w_i < u_i$$

For people with AIDS at time of diagnosis, the following formula is used:

$$f(w_i|y_i^c, T_i = w_i) = \frac{f(y_i^c|w_i)\lambda_{aids}(w_i)S_{aids}(w_i)S_{diag}(w_i)f(w_i)}{\int_0^{u_i} f(y_i^c|w_i)\lambda_{aids}(w_i)S_{aids}(w_i)S_{diag}(w_i)f(w_i)dw_i}, 0 < w_i < u_i$$

where, for individual i, the majority of parameters are as described in (1):

- $w_i$ is the time between infection and diagnosis
- $T_i$ is a latent variable representing time from HIV diagnosis to AIDS onset
- $y_i^c$ is the CD4 cell count at time of diagnosis
- $f(y_i^c|w_i)$ is the likelihood of observing CD4 cell count $y_i^c$ at time $w_i$ since HIV infection. This likelihood differs according to individual characteristics like age, sex, and route of transmission.
- $\lambda_{aids}(w_i)$ is the hazard rate of developing AIDS at time $w_i$ since HIV infection. This hazard rate differs according to individual characteristics like age, sex, and route of transmission.
- $S_{aids}(w_i)$ is the survival function associated with $\lambda_{aids}(w_i)$, that is, the probability of not developing AIDS prior to time $w_i$ since HIV infection
- $u_i$ is the longest possible delay between infection and diagnosis, given the restrictions that no infections occur before 1980, before the age of 15, or before the date of the last negative test
- $f(w_i)$ is the prior distribution of observing time $w_i$ between HIV infection and diagnosis. This distribution is assumed to be uniformly distributed between 0.01 years and $u_i$

$S_{diag}(w_i)$, the survival function representing the probability of not being HIV diagnosed by the time $w_i$ since HIV infection, is newly added to our version of the model. The approach used to estimating this function is similar to that described in (2, 3). For repeat testers (people with a prior negative HIV test), we assume a uniform distribution. For people with no prior negative test, we assume testing prior to AIDS follows an exponential distribution, with parameters that may vary in strata defined by transmission route and place of birth:

$$S_{diag}(w_i) = \exp(-\lambda_{diag} w_i)$$

where

$$1/\lambda_{diag} = \frac{\text{scale}}{p_{aids}^{-(\frac{1}{\text{shape}})} - 1}$$

with the parameters defined as follows:

- *Shape* and *scale* are the parameters of a gamma distribution used to describe the distribution of the AIDS incubation period in the general population. We set *shape*=5.251, *scale*=1.974
- $p_{aids}$ is the proportion of HIV diagnoses at the AIDS stage for the stratum of interest

As this modification mainly affects people with long estimated delays between infection and diagnosis, it has only a minor impact of estimates of HIV incidence but leads to better estimates of the number of people with undiagnosed HIV infections.

**A.2 Derivation of a variance formula for the delay distributions between infection or migration and diagnosis**

As described in the main text, the conditional probability $p_{jk}$ is the probability of a diagnosis delay = $t_j$, given the delay is ≤$t_j$. These probabilities are modeled using a generalized linear model with a complementary log-log link:

$$\log(-log(1-p_{jk})) = \alpha_j + \beta X_k$$

where $X_k$ is a vector of covariates. We will refer to the vector of parameters of this model as $\gamma = (\alpha_1, \cdots, \alpha_m, \beta)^T$, where $\beta$ is a vector of regression coefficients.

Furthermore, the diagnosis delay distributions are:

$$F_k(t_s) = [F(t_s)]^{\theta_k}, \theta_k = \exp(\beta X_k), s = 0, \dots, m-1$$

where $F(t_s) = \prod_{i=s+1}^{m} \exp(-\exp(\alpha_i))$ is the diagnosis delay distribution when all of the covariates are 0.

The original Brookmeyer paper (4) provides a variance formula based on Greenwood's formula for the case in which all covariates are all categorical, and each stratum can be considered independently. Here we derive a more general variance formula that can, for example, allow the possibility of a linear relationship between $\log(-\log(1-p_{jk}))$ and the calendar year of infection.

Using the delta method, we obtain

$$\mathrm{var}\left(\hat{F}_k(t_s)\right) = \left[\hat{F}_k(t_s)\right]^2 \mathrm{var}\left(\log\left(\hat{F}_k(t_s)\right)\right)$$

Furthermore, we define

$$G(\gamma) = \log\left(\hat{F}_k(t_s)\right) = \hat{\theta}_k \log\left(\hat{\mathrm{F}}(t_s)\right) = -\left(\sum_{i=s+1}^{m} \exp(\hat{\alpha}_i)\right) \exp(\hat{\beta} X_k)$$

Applying the delta method again,

$$\mathrm{var}(\log\left(\hat{F}_k(t_s)\right) = \left(\frac{\partial G}{\partial \gamma}(\hat{\gamma})\right)^T V(\hat{\gamma}) \left(\frac{\partial G}{\partial \gamma}(\hat{\gamma})\right)$$

where $V(\hat{\gamma})$ is the variance-covariance matrix of the generalized linear model described above.

To simplify, we consider the case with only two covariates, such that $\theta_k = \exp(\beta_1 X_{k1} + \beta_2 X_{k2})$. Beginning with $t_0$, that is, a diagnosis delay of zero quarters:

$$\log\left(\hat{F}_k(t_0)\right) = -(\exp(\hat{\alpha}_1) + \cdots + \exp(\hat{\alpha}_m))\exp(\hat{\beta}_1 X_{k1} + \hat{\beta}_2 X_{k2})$$

$$\frac{\partial \log\left(\hat{F}_k(t_0)\right)}{\partial \alpha_1} = -\exp(\hat{\alpha}_1 + \hat{\beta}_1 X_{k1} + \hat{\beta}_2 X_{k2}) \coloneqq a_1$$

$$\vdots$$

$$\frac{\partial \log\left(\hat{F}_k(t_0)\right)}{\partial \alpha_m} = -\exp(\hat{\alpha}_m + \hat{\beta}_1 X_{k1} + \hat{\beta}_2 X_{k2}) \coloneqq a_m$$

$$\frac{\partial \log\left(\hat{F}_k(t_0)\right)}{\partial \beta_1} = -X_{k1}\left(\sum_{i=1}^{m} \exp(\hat{\alpha}_i)\right)\exp\left(\hat{\beta}_1 X_{k1} + \hat{\beta}_2 X_{k2}\right) = X_{k1}\sum_{i=1}^{m} a_i \coloneqq a_{m+1}$$

$$\frac{\partial \log\left(\hat{F}_k(t_0)\right)}{\partial \beta_2} = -X_{k2}\left(\sum_{i=1}^{m} \exp(\hat{\alpha}_i)\right)\exp\left(\hat{\beta}_1 X_{k1} + \hat{\beta}_2 X_{k2}\right) = X_{k2}\sum_{i=1}^{m} a_i \coloneqq a_{m+2}$$

To summarize,

$$\left(\frac{\partial G}{\partial \gamma}(\hat{\gamma})\right) = \begin{pmatrix} a_1 \\ \cdots \\ a_m \\ a_{m+1} \\ a_{m+2} \end{pmatrix}$$

And therefore, referring to the elements in $V$ as $v_{ij}$

$$\operatorname{var}(\log\left(\hat{F}_k(t_0)\right) = \sum_{i=1}^{m+2}\sum_{j=1}^{m+2} v_{ij} a_i a_j$$

Next considering $t_{m-1}$, that is, a diagnosis delay of just under the maximum allowed delay:

$$\log\left(\hat{F}_k(t_{m-1})\right) = -\left(\exp(\hat{\alpha}_m)\right)\exp\left(\hat{\beta}_1 X_{k1} + \hat{\beta}_2 X_{k2}\right)$$

$$\frac{\partial \log\left(\hat{F}_k(t_{m-1})\right)}{\partial \alpha_1} = \cdots = \frac{\partial \log\left(\hat{F}_k(t_{m-1})\right)}{\partial \alpha_{m-1}} = 0$$

$$\frac{\partial \log\left(\hat{F}_k(t_{m-1})\right)}{\partial \alpha_m} = -\exp\left(\hat{\alpha}_m + \hat{\beta}_1 X_{k1} + \hat{\beta}_2 X_{k2}\right) \coloneqq a_m$$

$$\frac{\partial \log\left(\hat{F}_k(t_{m-1})\right)}{\partial \beta_1} = -X_{k1}\exp\left(\hat{\alpha}_m + \hat{\beta}_1 X_{k1} + \hat{\beta}_2 X_{k2}\right) \coloneqq a_{m+1}$$

$$\frac{\partial \log\left(\hat{F}_k(t_{m-1})\right)}{\partial \beta_2} = -X_{k2}\exp\left(\hat{\alpha}_m + \hat{\beta}_1 X_{k1} + \hat{\beta}_2 X_{k2}\right) \coloneqq a_{m+2}$$

$$\operatorname{var}(\log\left(\hat{F}_k(t_{m-1})\right) = \sum_{i=m}^{m+2}\sum_{j=m}^{m+2} v_{ij} a_i a_j$$

The general formula can thus be expressed as:

$$\operatorname{var}(\log\left(\hat{F}_k(t_s)\right) = \sum_{i=s+1}^{m+2}\sum_{j=s+1}^{m+2} v_{ij} a_i a_j$$

**A.3 Approximately log-linear decline in old infections/migrations**

<u>People infected in the country of interest</u>

Yearly diagnoses of people infected prior to 2012: observed (red) versus predicted by our log-linear model of decline (blue). Results are shown overall and by sub-population (MSM born abroad and MSM born in France). Results from one infection time draw on simulated data with a drop in diagnoses from 2020-21 (the "COVID" dataset).

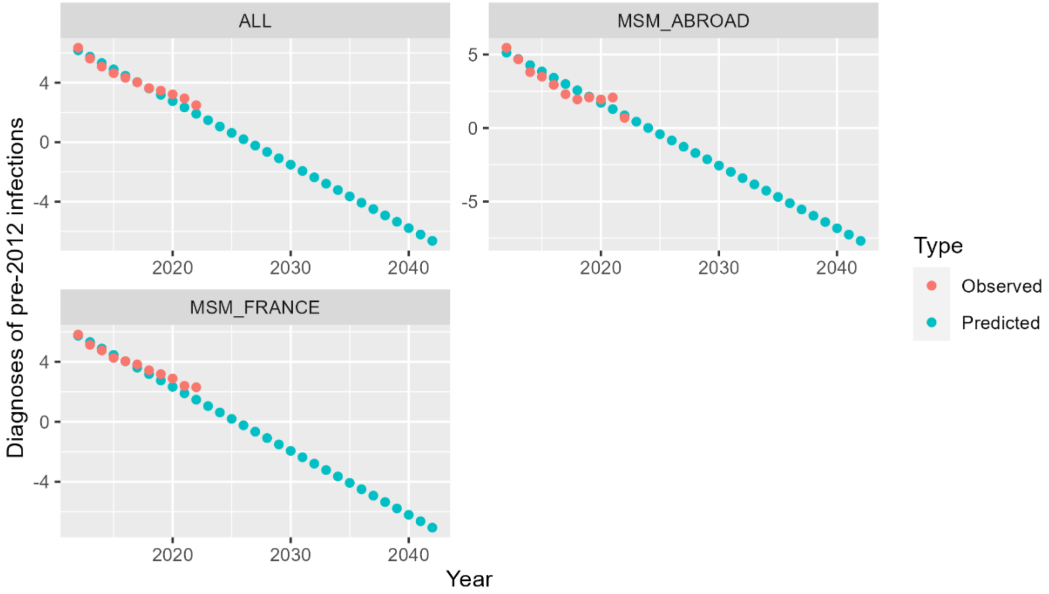


Yearly diagnoses of people infected prior to 2012: observed (red) versus predicted by our log-linear model of decline (blue). Results are shown overall and by sub-population (MSM born abroad and MSM born in France). Results from one infection time draw on simulated data with no drop in diagnosis probabilities in 2020-21.

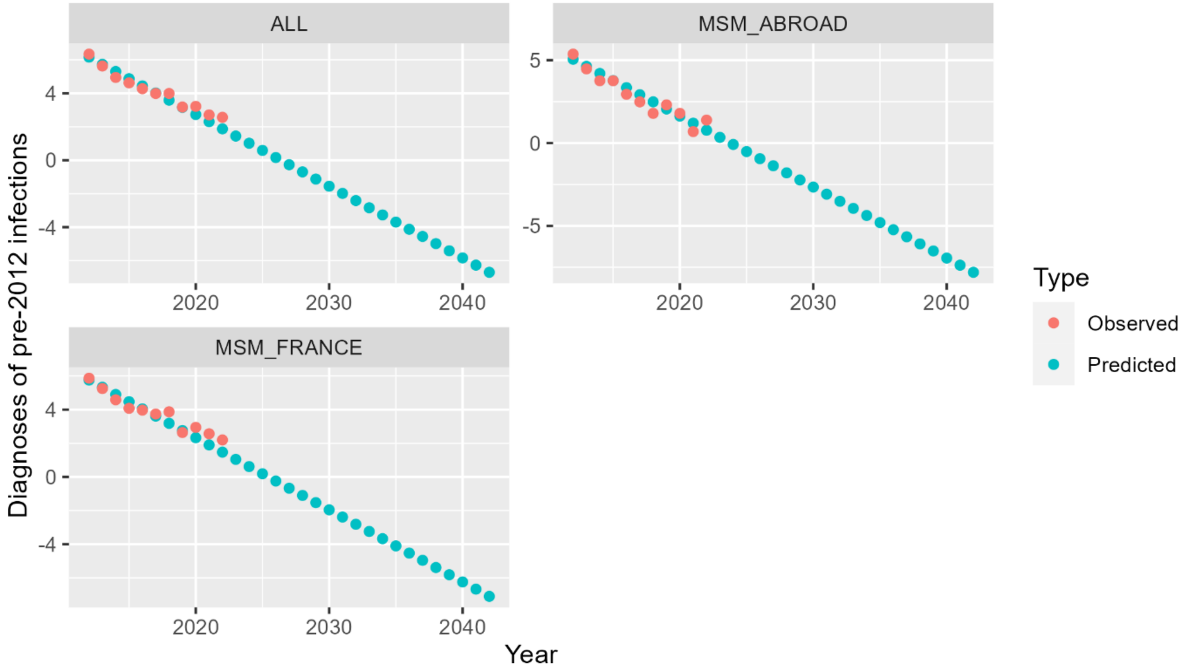

People with pre-migration infections

Yearly diagnoses of people who immigrated prior to 2012 with pre-migration infections: observed (red) versus predicted by our log-linear model of decline (blue) for MSM born abroad. Results from one infection time draw on simulated data with a drop in diagnosis probabilities in 2020-21 (the "COVID" dataset).

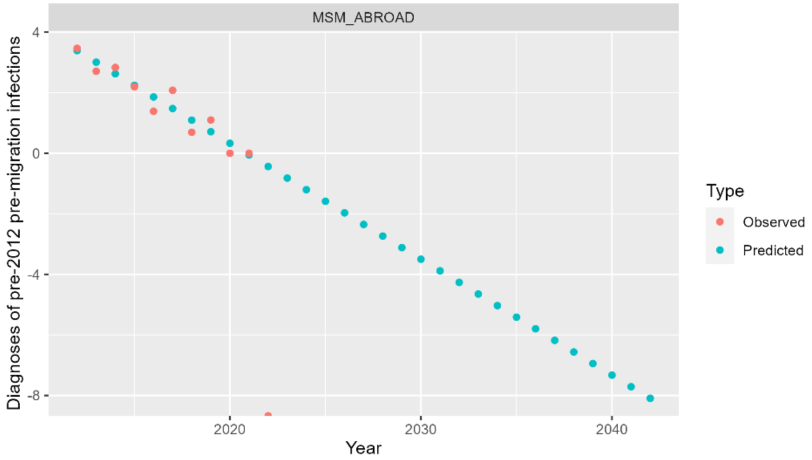


Yearly diagnoses of people who immigrated prior to 2012 with pre-migration infections: observed (red) versus predicted by our log-linear model of decline (blue) for MSM born abroad. Results from one infection time draw on simulated data with no drop in diagnosis probabilities in 2020-21.

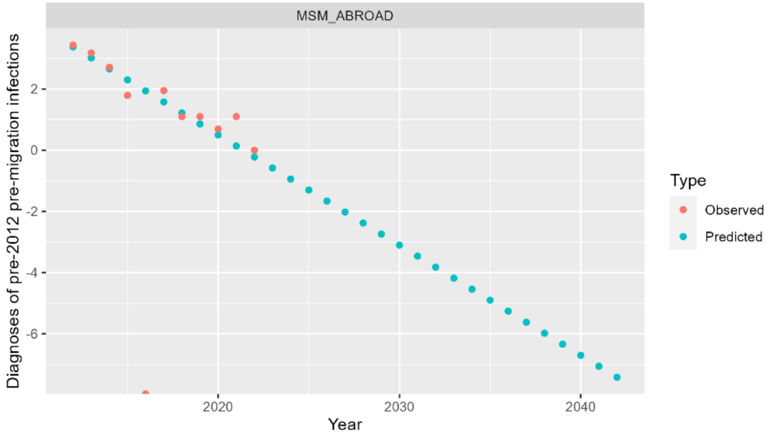

### A.4 Detailed description of simulated dataset

We used a simulated dataset, described here, to evaluate the performance of the model. The simulated data were inspired by previous simulated datasets of HIV cases including the HIV Synthesis model (5-8) and the simulated data described in (9). Parameters were modified for better alignment with French HIV surveillance data.

Incidence and infection dates

We assumed that HIV infection first began in a given European Country A in 1982, and that the number of new infections peaked in 1987 with 15,000 cases. We assumed a linear increase between 1982 and 1987. After 1987, we assumed a linear decrease in incidence until 2004. From 2004-2018, we assumed yearly incidence followed estimates produced by (10). Finally, we assumed the number of new infections was stable from 2018-2022. Within each year of infection, dates of infection were chosen according to a uniform distribution.

Patient characteristics

To facilitate alignment with the HIV Synthesis model, all simulated individuals were assumed to have men who have sex with men (MSM) born in Europe. Age at the time of infection was simulated according to a Normal(28,10) distribution, with a minimum age of 16 years (otherwise, a new age is drawn). We assumed 50% of individuals in the dataset were born outside of Country A.

Viral load and CD4

Following the HIV Synthesis model, we simulated individual viral load and CD4 trajectories over time (in quarters) for each new infection. Viral load was defined on a log10 scale. The log viral load at time of infection, V(0), was drawn according to a Normal(4,0.5) distribution truncated at 6.5. For each subsequent quarter, the change in viral load from the previous quarter was drawn according to a Normal(0.02275,0.05) distribution, again with a maximum value of 6.5 at each point in time. These distribution represent variations in the true underlying viral load; to account for measurement error and day-to-day variability in viral load measures, the "measured" log10 viral load at any given time point was assumed to equal the "true" log10 viral load at that time, plus a random value drawn from a Normal(0,0.2) distribution.

The baseline CD4 value at time of infection was assumed to depend on the viral load, with CD4 defined on a square-root scale. At the point of infection, the square root of the CD4 was assumed to be

$$Csq(0) = 32 - 2V(0) + N(0,2)$$

With a minimum value of 324 and a maximum value of 1500. The change in CD4 at each subsequent quarter was assumed to depend on the individual's age and viral load.

$$Csq(t) = Csq(t-1) + \Delta Csq(t-1)$$

$$\Delta Csq(t-1) \sim N(\gamma + \alpha, 1.2)$$

With γ and α defined as follows:

| Log10 viral load: V(t-1) | γ |
|---|---|
| <3 | -0.027 |
| [3,3.5) | -0.072 |

| Age(t-1) | α |
|---|---|
| <20 | 0.15 |
| [20,25) | 0.09 |

| | |
|---|---|
| [3.5,4) | -0.135 |
| [4,4.5) | -0.180 |
| [4.5,5) | -0.45 |
| [5,5.5) | -0.9 |
| [5.5,6) | -1.8 |
| ≥6 | -2.25 |

| | |
|---|---|
| [25,30) | 0.06 |
| [30,35) | 0 |
| [35,40) | 0 |
| [40,45) | -0.06 |
| [45,50) | -0.09 |
| [50,60) | -0.15 |
| ≥60 | -0.2 |

Older individuals thus tend to experience more rapid drops in CD4 count over time. CD4 was assumed to always remain ≥0 with no set maximum. As for viral load, these distributions represent variations in the true underlying CD4 value; to account for measurement error and day-to-day variability, the "measured" square root of CD4 at any given time point was assumed to equal the "true" square root of CD4, plus a random value drawn from a Normal(0,2) distribution. To account for the temporary fall in CD4 values in primary HIV infection, a temporary fall in the measured CD4 values at time 0 was modeled using a Normal(-150,50) distribution.

<u>Symptoms and AIDS defining conditions</u>

Again following the HIV Synthesis model, simulated individuals develop AIDS defining characteristics at an annual rate β that depends on CD4, viral load, and age. The probability of developing an AIDS-defining condition between time points (quarters) is then calculated as $1 - \exp(-0.25\beta)$.

In the HIV Synthesis model, the rate β is defined as:

$$\beta = \delta\theta \left(\frac{\text{age}}{38}\right)^{1.2}$$

Where δ and θ are as follows:

| CD4 | δ |
|---|---|
| ≥650 | 0.002 |
| [500,650) | 0.01 |
| [450,500) | 0.013 |
| [400,450) | 0.016 |
| [375,400) | 0.02 |
| [350,375) | 0.022 |
| [325,350) | 0.025 |
| [300,325) | 0.03 |
| [275,300) | 0.0375 |

| CD4 | δ |
|---|---|
| [250,275) | 0.045 |
| [225,250) | 0.055 |
| [200,225) | 0.065 |
| [175,200) | 0.08 |
| [150,175) | 0.1 |
| [125,150) | 0.13 |
| [100,125) | 0.17 |
| [90,100) | 0.2 |
| [80,90) | 0.23 |

| CD4 | δ |
|---|---|
| [70,80) | 0.28 |
| [60,70) | 0.32 |
| [50,60) | 0.4 |
| [40,50) | 0.5 |
| [30,40) | 0.8 |
| [20,30) | 1.1 |
| [10,20) | 1.8 |
| [0,10) | 2.5 |

| Log10 viral load: V(t) | θ |
|---|---|
| <3 | 0.2 |
| [3,4) | 0.3 |
| [4,4.5) | 0.6 |
| [4.5,5) | 0.9 |
| [5,5.5) | 1.2 |
| ≥5.5 | 1.6 |

To obtain a proportion of individuals newly diagnosed with HIV at the AIDS stage of infection more similar to that observed in French surveillance data, we modified the rate δ by adding a value of 0.025 regardless of CD4, viral load, or age:

$$\beta = (\delta + 0.025)\theta\left(\frac{\text{age}}{38}\right)^{1.2}$$

Within the French surveillance data, newly diagnosed cases who have symptoms of HIV infection, but who do not have AIDS-defining conditions or evidence or primary infection, are classified as "symptomatic non-AIDS cases". For our simulated dataset, we assumed that non-AIDS symptoms appear at a rate 8x that of AIDS-defining conditions (vs 5x for the apparition of CDC Category B symptoms in the HIVSynthesis model). We also assumed that 10% (for repeat testers) or 20% (for first-time testers) of these conditions are pulmonary tuberculosis, which are accordingly classed as AIDS cases. For ease of modeling and better concordance with French surveillance data, disease states were assumed to be transitory for first-time testers (lasting 3 months each), and absorbing for repeat testers.

Diagnosis: first-time testers

Among first-time testers, the quarter of diagnosis is determined according to probabilities that vary according to clinical stage. The date of diagnosis is then drawn from a uniform distribution within that quarter.

The annual rate of diagnosis in asymptomatic individuals, D, varies by calendar year. There are assumed to be no diagnoses of asymptomatic individuals prior to 1984. A high "catch-up" rate of testing (0.28) is applied from 1984-1985. Following that, annual rates of diagnosis are assumed to be stable at 0.12 from 1986-2001 before increasing linearly from 2001-2012 and remaining fixed at a rate of 0.164 from 2012 onwards. These values were selected from the mean values of the distributions used in the HIV Synthesis model. Annual rates are converted into a probability of diagnosis per quarter using the expression $1 - \exp(-0.25D)$.

We consider the first three months of HIV infection to be "primary HIV infection." Prior to 1995, the probability of diagnosis for individuals in primary infection was assumed to be the same as those with asymptomatic infections. Beginning in 1995, the probability of diagnosis in primary infection was set to 0.11. This value was modified from its original value of 0.2 in the HIV Synthesis model, to allow a better fit to the proportion of new HIV diagnoses in primary infection in French surveillance data.

For individuals with AIDS-defining conditions, excluding pulmonary tuberculosis, the probability of diagnosis before the next quarter is assumed to be 0.9, as in the HIV Synthesis model. For individuals with pulmonary tuberculosis and no other AIDS-defining conditions, this probability is assumed to be 0.42 (vs 0.5 in the original model). For symptomatic individuals without any AIDS-defining conditions, we assume the probability of diagnosis before the next quarter is 0.2 (vs 0.15 in the original model).

Diagnosis: repeat testers

HIV testing in individuals with a previous negative test is assumed to follow a Poisson process, similar to (9). In contrast to first-time testers, a date of diagnosis is thus determined for each individual prior to simulating his trajectories of viral load, CD4 count, and symptoms.

First, a date of first negative test is selected according to a uniform distribution in the 3 years prior to infection. The delays between tests are assumed to follow an exponential distribution with parameter λ that produces similar delays between the last negative test and the first positive test as those observed in the French surveillance data (a mean of approximately 28 months). Because the length of these intervals is conditional on the infection occurring between them, we assume that the time between last negative test and infection and the time between infection and first positive test both follow an exponential distribution with parameter λ, and that the delay between last negative test and first positive test thus follows a Gamma(2, λ) distribution. We used the R package gamlss (11) to estimate λ and thus arrived at a value of approximately 1/(12.75 months). Starting at the date of first negative test, subsequent tests are thus simulated according to an Exponential(1/12.75) distribution. The date of the first test that occurs after infection is taken as the date of diagnosis.

CD4, viral load, and symptoms at the time of diagnosis are then determined based on the trajectories described above. Diagnoses occurring within one quarter of infection are considered to be in primary infection. Initial simulations under-estimated the probability of symptoms or AIDS-defining conditions in repeat testers, as these states do not directly lead to diagnosis. To obtain a distribution of diagnoses by clinical stage more consistent with French surveillance data, symptoms and AIDS-defining conditions in repeat testers were assumed to be absorbing states, as previously described.

<u>COVID-19 pandemic related drop in diagnoses</u>

A decrease in the number of diagnosed new HIV cases was observed in France from 2020-2021, coinciding with the COVID pandemic. We were concerned that this could represent a decrease in the probability of diagnosis during those years, and wished to test the ability of the method to account for such a drop. We thus created two versions of the simulated dataset, a baseline dataset with no COVID-related drop in diagnoses in 2020 and 2021, and a "COVID" dataset containing such a drop.

For first-time testers in the "COVID" dataset, we multiplied the annual rate of diagnosis in asymptomatic infection by 1/2 in 2020 and 2/3 in 2022 compared to its baseline value. We similarly multiplied the probability of diagnosis in primary infection by 1/2 in 2020 and 2/3 in 2021. For repeat testers, the clinical stage is determined subsequently to the date of diagnosis. For simplicity, we therefore assumed the COVID pandemic only affected diagnoses during primary infection (i.e. the first three months following infection). Diagnoses initially selected to fall during this period were assumed to still take place with a probability of 1/2 (in 2020) and 2/3 (in 2021). Otherwise, a new date of diagnosis was simulated according to the same Poisson process described above, starting from the date of the missed potential diagnosis.

<u>Arrival in France</u>

Dates of arrival in Country A were simulated for the 50% of individuals in the dataset assumed to be born outside the country. We aimed for the distribution of the time between arrival in Country A and HIV diagnosis in the simulated data to be similar to that observed for MSM born in other European countries in the French surveillance data.

An exponential distribution was initially used for this distribution. However, the proportion of individuals initially diagnosed in the same year of their arrival in France was much higher in the surveillance data than could be explained with this distribution. We therefore used a zero-inflated model, in which a certain proportion of people were estimated to be diagnosed the same year as their arrival (with a day chosen uniformly between January 1 and the day of diagnosis) and the remainder diagnosed following an exponential distribution of delays. These two parameters were estimated using a zero-inflated Poisson regression model from the R package pscl (12).

<u>Sample</u>

The final size of the complete simulated dataset was 338,652 rows. For ease of use, we took a random sample of 50,000 of these rows, approximately 15% of the original dataset, to test the performance of the model described in this article.

<u>Comparisons of French surveillance data of new HIV diagnoses and the simulated dataset</u>

The plots below compare the simulated dataset with the French mandatory declaration dataset of new HIV diagnoses. Both a raw surveillance dataset and a dataset with missing values addressed by multiple imputation are shown. As the simulated dataset represents only MSM born in Country A or other European countries, over 15 years old and diagnosed from 2012-2022, the same filters are applied to the surveillance data for all comparisons.

Percentage of new diagnoses by clinical stage (AIDS, Asymptomatic, Primary HIV infection, or Symptomatic non-AIDS), testing history (repeat or first-time tester) and year. The simulation data is very similar to the French surveillance data on new HIV diagnoses.

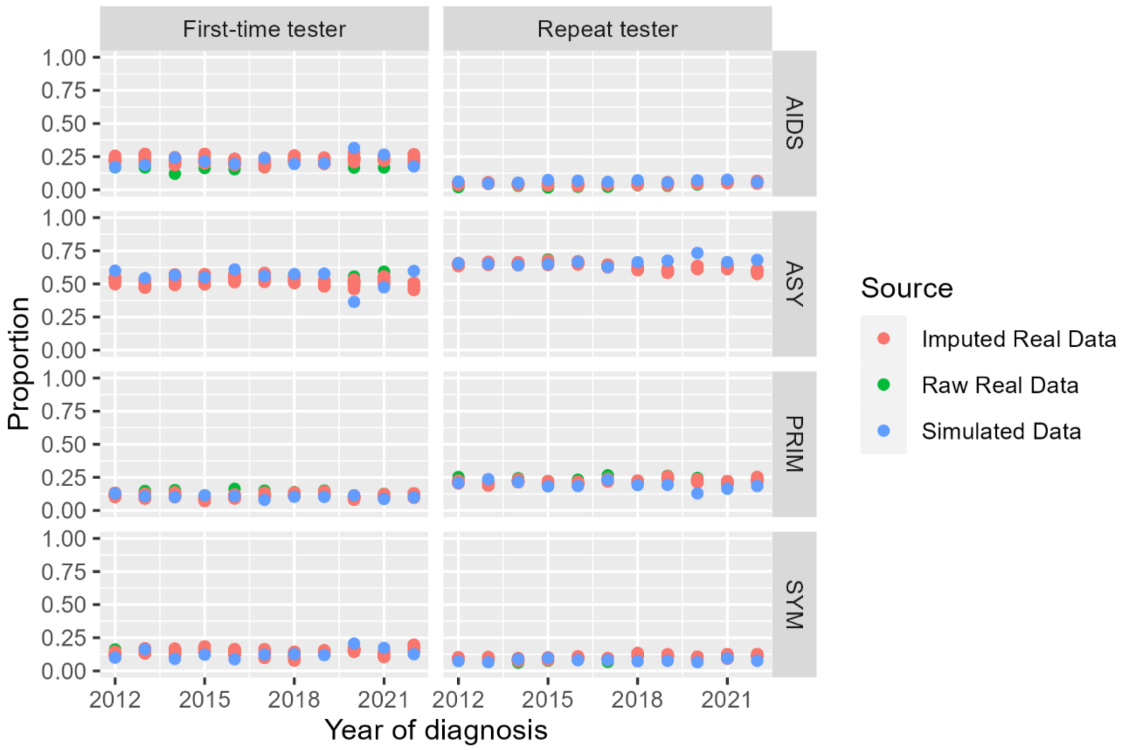

Category of CD4 cell count at HIV diagnosis by clinical stage and testing history. The CD4 cell counts in the simulated dataset are similar to those observed in the French surveillance data for people in primary stage, asymptomatic, or symptomatic non-AIDS stages of infection. For individuals at AIDS stage, CD4 counts in the simulated dataset tend to be higher than those in the surveillance dataset, particularly among repeat testers. Because the time from last negative test and first positive test is often short, it was difficult to obtain a realistic proportion of repeat testers at AIDS stage without accepting these higher CD4 values.

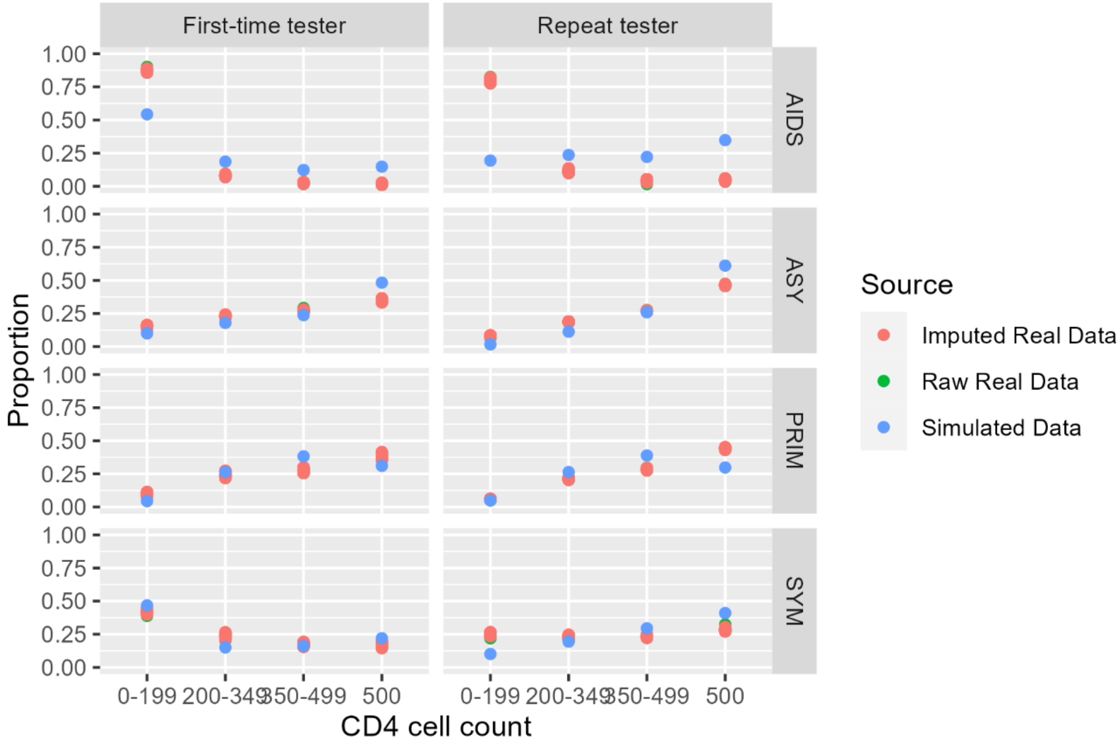

Age at HIV diagnosis by clinical stage and testing history. The distribution of ages was not modified from the original HIV Synthesis model despite discordance with the French data, as this variable has limited impact on the models used in this paper.

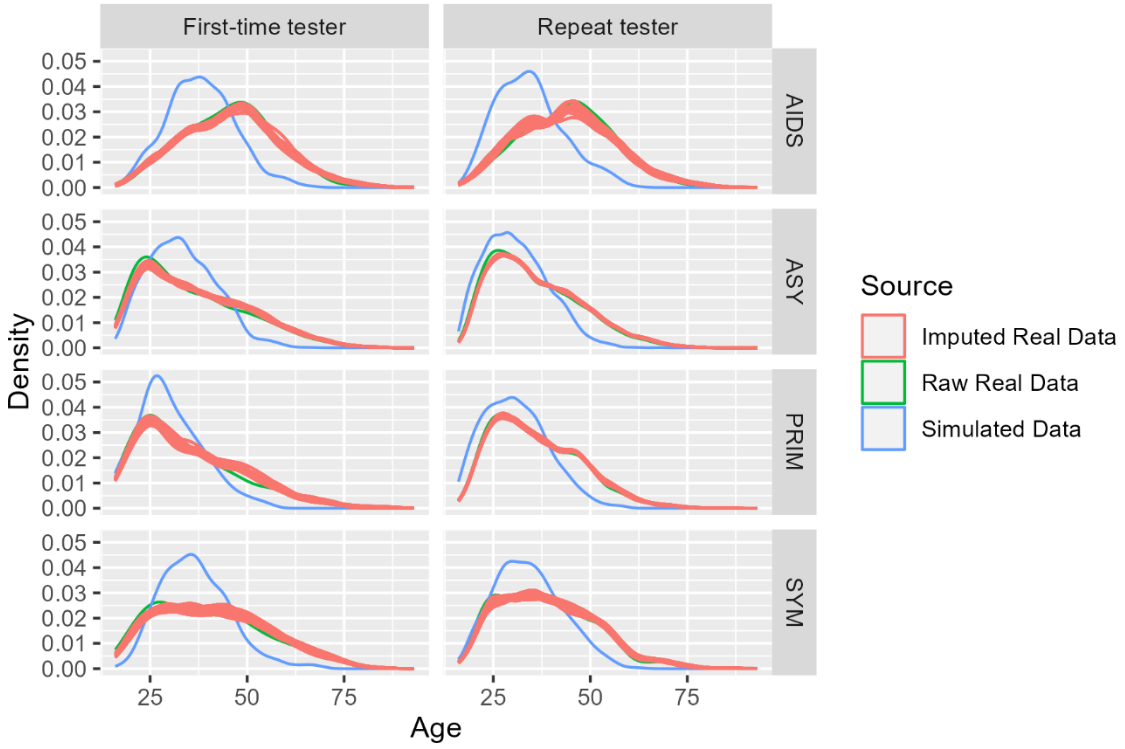

Viral load at HIV diagnosis by clinical stage and testing history. The simulated viral loads tend to be somewhat lower than the values observed in the French surveillance data on new diagnoses, particularly for individuals in primary HIV infection.

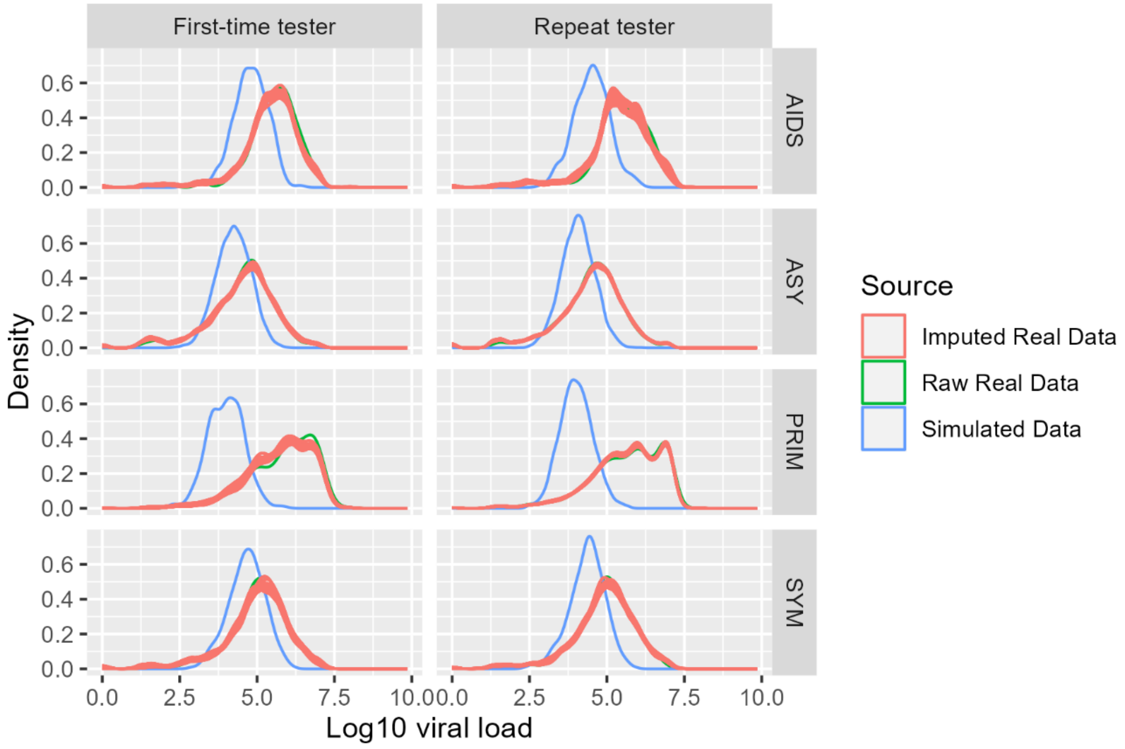


Type of tester (repeat or first-time) by clinical stage at diagnosis. Good coherence is observed between the simulated and real French surveillance data.

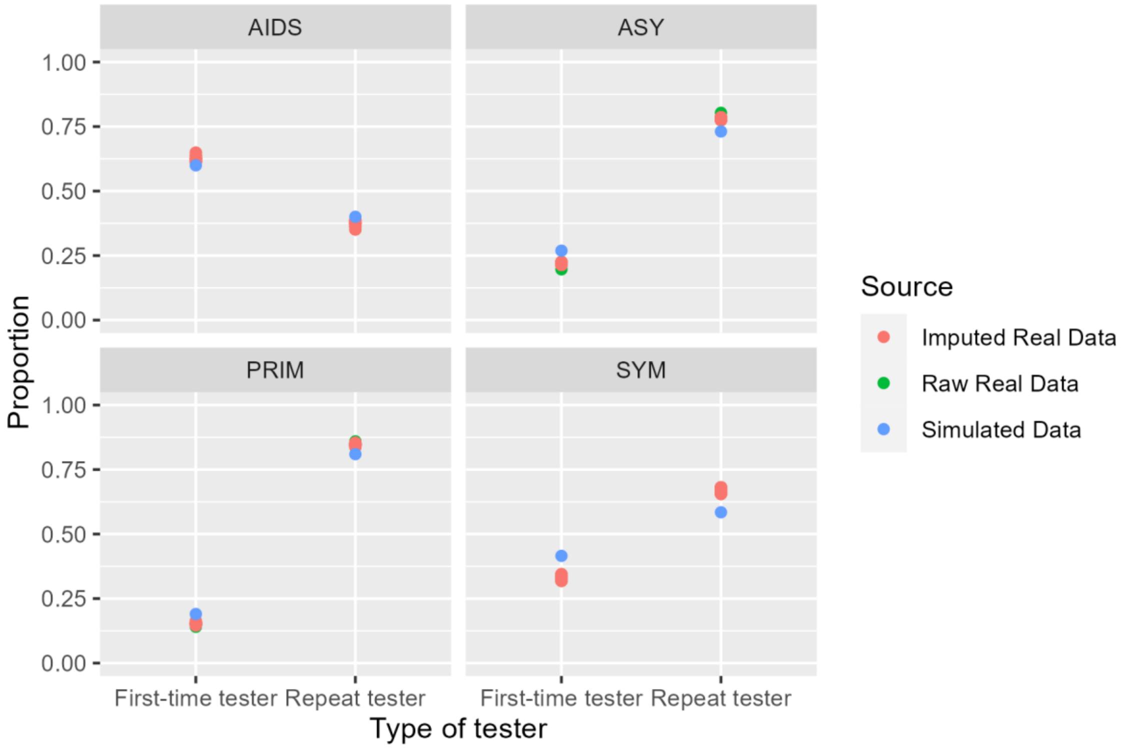

Delay (in months) between last negative and first positive HIV test by clinical stage, among repeat testers. Despite some differences between the two datasets, a similar trend is observed towards more recent tests among people in primary stage of infection and longer delays since last negative test among people in AIDS stage.

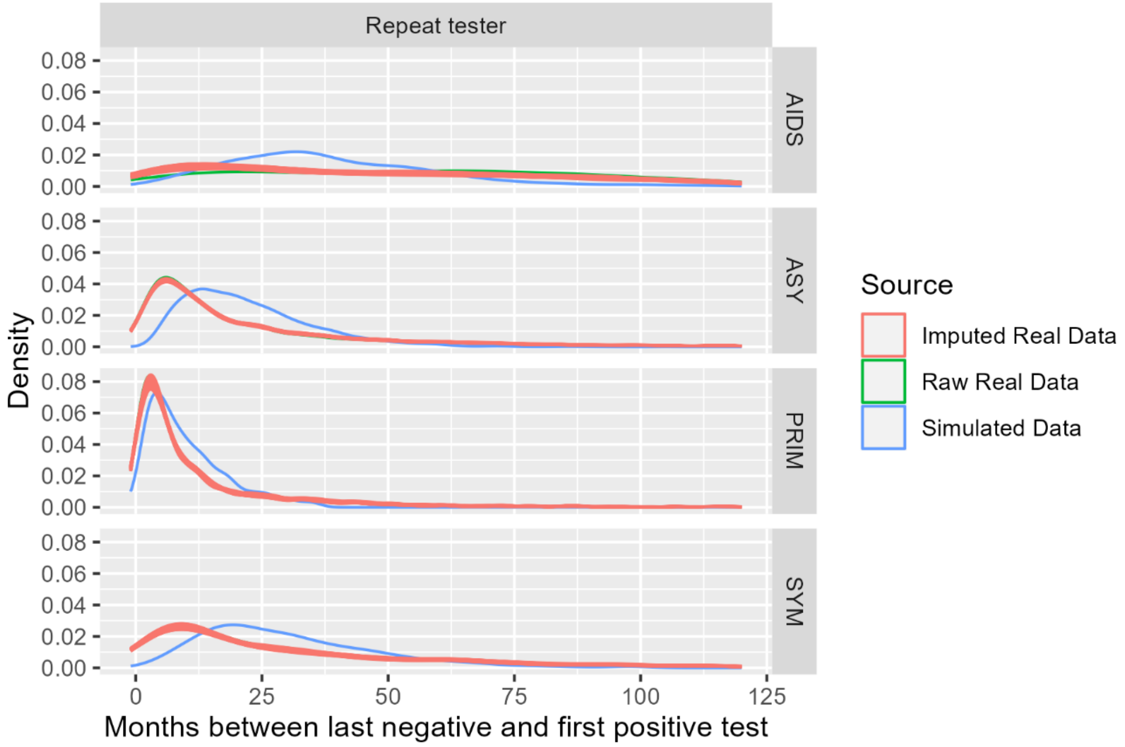

Delay (in years) between last negative and first positive HIV test by clinical stage, among people born abroad. Because relatively few MSM diagnosed with HIV in France were born in other European countries, there is considerable uncertainty in the imputed delays from the real data. Nevertheless the distributions are relatively similar between simulated and real data, with a notable spike in diagnoses in the first five years following diagnosis.

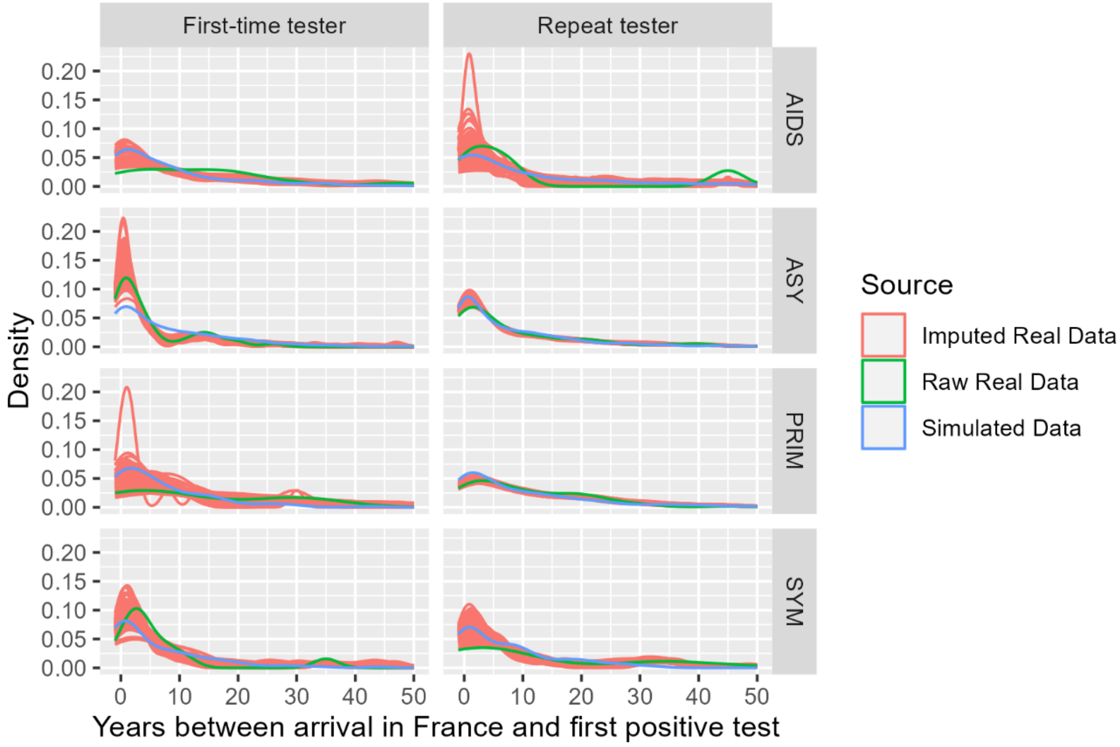
