## Supplemental Data and Programmes for "A new approach to estimating HIV incidence and the size of the infected and undiagnosed population in high immigration settings": Estimate_Incidence_Undiag_Final.R

```
# Part II: Estimate incidence, arrival of undiagnosed HIV+ migrants, and the
# number of undiagnosed infections
# Using output of Estimate_Infection_Dates_Final.R


rm(list=ls())
gc(reset=T)

library(tidyverse)
library(openxlsx)
library(lubridate)
library(mvtnorm)
library(splines)
library(purrr)
library(foreach)
library(mice)
library(MASS,exclude=c('select'))
library(car)

# Make estimates with correct accounting for migration, or 
# ignoring migration?
AccountForMig = T # T F

# COVID drop in diagnoses version
# sim_base_full <- 'bdd_testeurs_covid_mig.Rds'
# est_inf_dates <- 'sim_output_fast_covid.Rds'
# PATH <- './Covid/'

# # No COVID drop in diagnoses version (use one or the other)
sim_base_full <- 'bdd_testeurs_mig.Rds'
est_inf_dates <- 'sim_output_fast_nocovid.Rds'
PATH <- './Nocovid/'

########## Read simulated data for comparison with results

don_samp <- readRDS(sim_base_full)

don_samp <- don_samp %>%
  mutate(mig.inf.true = if_else(date.arriv>date.infection,"PreMig","PostMig"),
         place.inf.true = if_else(pnais=="FRANCE"|mig.inf.true=="PostMig","FRANCE","OTHER")) %>%
  # place of birth and transmission route in 1 variable
  mutate(mcont_pnais = if_else(pnais=="FRANCE"&mcont=="HOMO-BI", "MSM_FRANCE", "MSM_ABROAD"),
         mcont_all = mcont_pnais)
  
# True number undiagnosed at the end of 2022
# INFECTED IN FRANCE
true_undiag_2022_france <- don_samp %>%
  filter(annee.infection<2023 & annee.diagnostic>2022 &
           (place.inf.true=="FRANCE")) %>%
  group_by(annee.infection,mcont_all) %>%
  summarise(Undiag=n())
true_undiag_2022_france <- true_undiag_2022_france %>%
  bind_rows(true_undiag_2022_france %>% group_by(annee.infection) %>%
              summarise(Undiag=sum(Undiag)) %>%
              mutate(mcont_all="ALL"))

# True number undiagnosed at the end of 2022
# INFECTED ABROAD
true_undiag_2022_abroad <- don_samp %>%
  filter(annee.infection<2023 & annee.diagnostic>2022 &
           (place.inf.true!="FRANCE"&annee.arriv<2023)) %>%
  group_by(annee.migration=annee.arriv,mcont_all) %>%
  summarise(Undiag=n())
true_undiag_2022_abroad <- true_undiag_2022_abroad %>%
  bind_rows(true_undiag_2022_abroad %>% group_by(annee.migration) %>%
              summarise(Undiag=sum(Undiag)) %>%
              mutate(mcont_all="ALL"))

# True number undiagnosed at the end of 2022
# IGNORING MIGRATION/ AS IF ALL INFECTED IN FRANCE
true_undiag_2022_nomig <- don_samp %>%
  filter(annee.infection<2023 & annee.diagnostic>2022) %>%
  group_by(annee.infection,mcont_all) %>%
  summarise(Undiag=n())
true_undiag_2022_nomig <- true_undiag_2022_nomig %>%
  bind_rows(true_undiag_2022_nomig %>% group_by(annee.infection) %>%
              summarise(Undiag=sum(Undiag)) %>%
              mutate(mcont_all="ALL"))
true_undiag_2022_nomig_overall <- true_undiag_2022_nomig %>%
  group_by(mcont_all) %>%
  summarise(N_undiag = sum(Undiag)) %>%
  mutate(Assumption = 'NoMig')

# True number undiagnosed at the end of 2022
# TOTAL - by year of entry (via migration or infection)
true_undiag_2022 <- don_samp %>%
  mutate(annee.entry = if_else(place.inf.true=="FRANCE",annee.infection,annee.arriv)) %>%
  filter(annee.infection<2023 & annee.diagnostic>2022 &
           (place.inf.true=="FRANCE"|date.arriv<2023)) %>%
   group_by(annee.entry,mcont_all) %>%
  summarise(Undiag=n())
true_undiag_2022 <- true_undiag_2022 %>%
  bind_rows(true_undiag_2022 %>% group_by(annee.entry) %>%
              summarise(Undiag=sum(Undiag)) %>%
              mutate(mcont_all="ALL"))
true_undiag_2022_overall <- true_undiag_2022 %>%
  group_by(mcont_all) %>%
  summarise(N_undiag = sum(Undiag)) %>%
  mutate(Assumption = 'TrueMig') 

# Save true undiagnosed 
wb <- createWorkbook()
addWorksheet(wb,"TrueMig")
addWorksheet(wb,"NoMig")
writeData(wb,sheet="TrueMig",true_undiag_2022_overall)
writeData(wb,sheet="NoMig",true_undiag_2022_nomig_overall)
saveWorkbook(wb = wb, file = paste0(PATH,"/True_Sim_Undiag.xlsx"), overwrite = T)  


##############################
#### Record true incidence values in France
#### And true # arrivals/year
##############################

# Incidence, if all infected in France
true_inc_nomig <- don_samp %>%
  group_by(annee.infection,mcont_all) %>%
  summarise(Inc = n()) %>%
  mutate(Source = 'True')
diag_dat <- don_samp %>%
  group_by(annee.diagnostic,mcont_all) %>%
  summarise(Inc = n()) %>%
  mutate(Source = 'Diagnoses') %>%
  mutate(annee.infection=annee.diagnostic)
true_inc_nomig <- bind_rows(true_inc_nomig,diag_dat)
true_inc_nomig <- true_inc_nomig %>%
  bind_rows(true_inc_nomig %>%
              group_by(annee.infection,Source) %>%
              summarise(Inc=sum(Inc)) %>%
              mutate(mcont_all="ALL"))

true_inc <- don_samp %>%
 filter(place.inf.true=="FRANCE") %>%
  group_by(annee.infection,mcont_all) %>%
  summarise(Inc = n()) %>%
  mutate(Source = 'True')
diag_dat <- don_samp %>%
  filter(place.inf.true=="FRANCE") %>%
  group_by(annee.diagnostic,mcont_all) %>%
  summarise(Inc = n()) %>%
  mutate(Source = 'Diagnoses') %>%
  mutate(annee.infection=annee.diagnostic)
true_inc <- bind_rows(true_inc,diag_dat)
true_inc <- true_inc %>%
  bind_rows(true_inc %>%
              group_by(annee.infection,Source) %>%
              summarise(Inc=sum(Inc)) %>%
              mutate(mcont_all="ALL"))

true_arriv <- don_samp %>%
  filter(place.inf.true=="OTHER") %>%
  group_by(annee.migration=annee.arriv,mcont_all) %>%
  summarise(Arriv = n()) %>%
  mutate(Source = 'True') %>%
  filter(annee.migration<=2022)
true_arriv <- true_arriv %>%
  bind_rows(true_arriv %>%
              group_by(annee.migration,Source) %>%
              summarise(Arriv=sum(Arriv)) %>%
              mutate(mcont_all="ALL"))


# Save true incidence/arrivals 
wb <- createWorkbook()
addWorksheet(wb,"IncTrueMig")
addWorksheet(wb,"ArrivTrueMig")
addWorksheet(wb,"IncNoMig")
writeData(wb,sheet="IncTrueMig",true_inc %>% filter(Source=='True') %>% 
            select(annee.infection,mcont_all,Inc))
writeData(wb,sheet="ArrivTrueMig",true_arriv %>% filter(Source=='True') %>% 
            select(annee.migration,mcont_all,Arriv))
writeData(wb,sheet="IncNoMig",true_inc_nomig %>% filter(Source=='True') %>% 
            select(annee.infection,mcont_all,Inc))
saveWorkbook(wb = wb, file = paste0(PATH,"/True_Sim_Inc_Mig.xlsx"), overwrite = T)  


#################################################################
#################################################################
################ Estimate HIV incidence in France
################ And number undiagnosed, infected in France
#################################################################
#################################################################

don_diag_full <- readRDS(est_inf_dates)


########## Organizing the data into grid format

don_diag_full <- don_diag_full %>%
  mutate(date.infection.est = date.diagnostic - dyears(DelayInfDiag)) %>%
  mutate(annee.infection.est = year(date.infection.est)) %>%
  mutate(true_delay = interval(date.infection,date.diagnostic)/years(1)) %>%
  mutate(q.inf = lubridate::quarter(date.infection.est,type="date_first"),
         q.diag = lubridate::quarter(date.diagnostic,type="date_first"),
         q.del = (month(q.diag) - month(q.inf))/3 + (year(q.diag)-year(q.inf))*4,
         a.inf = annee.infection.est, 
         a.diag = annee.diagnostic)  %>%
  mutate(mig.inf.est = if_else(date.arriv>date.infection.est,"PreMig","PostMig"),
         place.inf.est = if_else(pnais=="FRANCE"|mig.inf.est=="PostMig","FRANCE","OTHER")) %>%
  # place of birth and transmission route in 1 variable
  mutate(mcont_pnais = if_else(pnais=="FRANCE"&mcont=="HOMO-BI", "MSM_FRANCE", "MSM_ABROAD"),
         mcont_all = mcont_pnais)

# If want results of full model/accounting for migration
if (AccountForMig==T) {
  don_diag_full <- don_diag_full %>%
    # IMPORTANT: filter to infections in France
    filter(place.inf.est == "FRANCE") 
}         


# Data should only incude diagnoses through 2022
min_diag=as.Date("2012-01-01"); 
max_diag=as.Date("2022-12-31");
min_year_diag=year(min_diag)
max_quarter=lubridate::quarter(max_diag,type="date_first")
maxAn = year(max_diag) 

data <- don_diag_full %>%
  filter(annee.diagnostic<=2022)

kmax=10*4 # truncate delays at 10 years/40 quarters for incidence/migrations

don_inc_est <- NULL
coeffs <- NULL
pred_old <- NULL


############ Variance based on Delta method

n_imp <- max(data$.imp) # Imputations/infection time draws per individual in the data
#n_imp <- 5 # TO TEST ONLY
set.seed(100) # for reproducibility

for (i in 1:n_imp) {
  
  print(i)
  
  data_imp <- data %>% filter(.imp == i) 
  
  # Add an option estimating "ALL" transmission groups/regions of birth
  data_imp <- data_imp %>% 
    bind_rows(data_imp %>% mutate(mcont_all="ALL"))
  
  ##########################################################################
  #### Estimating diagnoses in people infected prior to min_year_diag
  
  old_inf <- data_imp %>% filter(a.diag>=min_year_diag) %>% 
    group_by(a.diag,mcont_all) %>% 
    summarise(n_diag=n(), n_old=sum(a.inf<min_year_diag)) 
  
  # Going to impose that number pre-2012 diagnosed after 2022 
  # Can't exceed 3*number diagnosed during 2012-2022 (seems reasonable given 
  # median time to aids diagnosis ~11 years)
  curr <- old_inf %>%
    group_by(mcont_all) %>%
    summarise(max_old=3*sum(n_old))
  
  # roughly log-linear decline
  ggplot(old_inf) +  geom_point(aes(x=a.diag,y=log(n_old)),col='red') +
    facet_wrap(vars(mcont_all))
  
  # fit Poisson model
  m_old <- glm(n_old~a.diag + mcont_all, 
               family=poisson(link="log"),
               data=old_inf)
  
  
  # transmission group categories, including "ALL"
  cat <- unique(data_imp$mcont_all)
  # dataset for predictions
  years <- sort(rep(seq(min_year_diag,min_year_diag+30),length(cat)))
  mcont <- rep(cat,length(seq(min_year_diag,min_year_diag+30)))
  pred_df <- bind_cols(a.diag=years,mcont_all=mcont)
  
  # To visually confirm how the model looks. 
  pred_df$n_old <- predict(m_old,pred_df,type="response")
  pred_df$Type <- 'Predicted'
  plot_df <- pred_df %>%
    bind_rows(old_inf %>% mutate(Type='Observed'))
  ggplot(plot_df) +
    geom_point(aes(x=a.diag,y=n_old,col=Type))+
    facet_wrap(vars(mcont_all),scales='free')
  ggplot(plot_df) +
    geom_point(aes(x=a.diag,y=log(n_old),col=Type))+
    facet_wrap(vars(mcont_all),scales='free',nrow=2) +
    ylab('Diagnoses of pre-2012 infections') + 
    xlab('Year')
  #ggsave('old_inf_plot.png',height=4,width=7,units='in')
  
  # Bootstraps for within-imputation variance
  pred_df <- pred_df %>% 
    filter(a.diag>=year(max_diag))
  pred_boot <- Boot(m_old, f=function(x)predict(x, pred_df, type="response"), R=100,
                    method="case")
  pred_boot <- bind_cols(pred_df,t(pred_boot$t))
  pred_boot_sum <- pred_boot %>%
    group_by(mcont_all) %>%
    summarise(across(starts_with('...'), ~sum(.)))
  # max value as specified above in case of small groups/unstable estimates
  pred_boot_sum$max_old <- curr$max_old 
  pred_boot_sum <- pred_boot_sum %>%
    mutate(across(starts_with('...'), ~if_else(.>max_old,max_old,.))) 
  # summarize bootstrap values
  pred_boot_sum$mean_diag <- apply(pred_boot_sum %>% select(starts_with('...')),1,function(x) mean(x,na.rm=T))
  pred_boot_sum$var_diag <- apply(pred_boot_sum %>% select(starts_with('...')),1,function(x) var(x,na.rm=T))
  pred_boot_sum <- pred_boot_sum %>% select(mcont_all,mean_diag,var_diag) %>%
    mutate(.imp=i)
  pred_old <- bind_rows(pred_old,pred_boot_sum)
  
  ####################################################################
  ## Estimating incidence and # undiagnosed for people infected in [min_diag,max_diag]
  
  data_imp <- data_imp %>%
    mutate(q.inf.full=q.inf,
           q.diag.full=q.diag,
           q.del.full=q.del,
           a.inf.full = a.inf, 
           a.diag.full = a.diag) %>%
    # Truncating for inc estimation only
    mutate(q.del = if_else(q.del>kmax, kmax, q.del),
           q.inf = q.diag - months(3*q.del),
           a.inf=year(q.inf)) %>%
    mutate(q.trunc.del = (month(max_quarter) - month(q.inf)) / 3 + 
             (year(max_quarter) - year(q.inf)) * 4) %>%
    filter(a.inf >= min_year_diag ) # needed for Brookmeyer
  
  # Start an inner loop here to estimate by route of transmission ('mcont')
  for (j in cat) {
    
    print(j)
    data_strat <- data_imp %>% filter(mcont_all==j)
    
    # Define regression formulas in advance
    
    # if all transmission groups, include variable mcont_pnais
    if (j=="ALL") { 
      vars_reg <- c('year_2020', 'year_cont', 'mcont_pnais') # Standard
      #vars_reg <- c('year_G20', 'year_cont_L20', 'mcont_pnais') # ECDC V1
      #vars_reg <- c('year_G20', 'year_cont_L20', 'year_cont_G20', 'mcont_pnais') # ECDC V2
      #vars_reg <- c('year_cont', 'mcont_pnais') # ECDC V3
    } else {
      vars_reg <- c('year_2020', 'year_cont') # Standard
      #vars_reg <- c('year_G20', 'year_cont_L20') # ECDC V1
      #vars_reg <- c('year_G20', 'year_cont_L20', 'year_cont_G20') # ECDC V2
      #vars_reg <- c('year_cont') # ECDC V3
    }
    
    reg_eqn <- paste(c("q.del",vars_reg),collapse="+") # for regression
    reg_form = as.formula(paste('Diag', "~0+", reg_eqn)) # want 0 here to have q.1 in output/no intercept
    
    mm_eqn <- paste(vars_reg,collapse="+") # for model matrix
    mm_form = as.formula(paste("~", mm_eqn)) # don't want 0+ here, messes up factors
    
    
    # if not at least 10 data points, with at least 2 different delays
    # Then error, insufficient data
    if (nrow(data_strat)<10 | length(unique(data_strat$q.del))<2) {
      stop("Too few data points to estimate incidence")
    }
    
    # Set first q.del = min(q.del) equal to zero quarters
    # Will usually be true; if not, for small strata, may have issues dividing by zero
    data_strat$q.del[which(data_strat$q.del==min(data_strat$q.del))[1]] <- 0
    
    
    # Formatting for GLM                                               
    # Will allow delay distributions to vary by year of infection (a.inf) 
    # in various forms, as well as by
    # mode of transmission/country of origin ("mcont_pnais")
    
    # First, creating grouped data
    
    # in case zeros
    inf_del_mat <- expand.grid(unique(data_imp$a.inf),
                               seq(0,kmax),unique(data_imp$mcont_pnais))
    colnames(inf_del_mat) <- c('a.inf','q.del','mcont_pnais') 
    
    data_grp <- data_strat %>%
      group_by(mcont_pnais, a.inf, q.del) %>%
      summarise(Yjk = n()) %>%
      full_join(inf_del_mat) %>%
      mutate(Yjk = replace_na(Yjk,0))
    
    # calculating njk (number at risk) by strata (a.inf) and delay
    # To calc nijk - first get number >=delay, then subtract those w/ trunc <delay
    # This is faster than doing in a loop
    num_Ldelay <- data_strat %>% # num w reporting delay <=q.del
      group_by(mcont_pnais,a.inf,q.del) %>% 
      summarise(n=n()) %>%
      full_join(inf_del_mat) %>%
      mutate(n = replace_na(n,0)) %>% 
      group_by(mcont_pnais,a.inf) %>%
      arrange(q.del) %>%
      mutate(n_q.del = cumsum(n)) 
    num_Ltrunc <- data_strat %>% # truncation time >= q.del
      group_by(mcont_pnais,a.inf,q.del=q.trunc.del) %>% 
      summarise(n=n()) %>%
      full_join(inf_del_mat) %>%
      mutate(n = replace_na(n,0)) %>% 
      group_by(mcont_pnais,a.inf) %>%
      arrange(q.del) %>%
      mutate(n_q.trunc = cumsum(n),
             n_q.trunc = n_q.trunc - n) %>% # for less than, not less than or equal
      select(-n) 
    num_all <- num_Ldelay %>%
      left_join(num_Ltrunc) %>%
      mutate(n_q.trunc=replace_na(n_q.trunc,0)) %>%
      mutate(njk=n_q.del-n_q.trunc) %>%
      select(a.inf,q.del,mcont_pnais,njk)
    data_grp <- data_grp %>%
      left_join(num_all)
    
    # estimating pjk and non_pos = njk-Yjk
    data_grp <- data_grp %>%
      mutate(
        year_2020 = if_else(a.inf == 2020, 1, 0),
        year_G20 = if_else(a.inf >= 2020, 1, 0),
        year_cont = a.inf - median(seq(min_year_diag,maxAn)),
        year_cont_L20 = if_else(a.inf < 2020, year_cont, 0),
        year_cont_G20 = if_else(a.inf >= 2020, year_cont, 0)
      )
    
    data_grp_full <- data_grp # incldues q.del=0, unlike below
    
    # estimating pjk and non_pos = njk-Yjk
    data_grp <- data_grp %>%
      mutate(pjk = Yjk / njk) %>%
      filter(njk > 0) %>%
      mutate(non_pos = njk - Yjk) %>% 
      filter(q.del != 0) %>% #because Yjk=njk
      ungroup() 
    
    # for regression, want 1 line per pos and 1 line per non-pos (per period at risk)
    don_reg_pos <- data_grp %>%
      uncount(Yjk) %>%
      select(a.inf, q.del, mcont_pnais, all_of(vars_reg)) %>% # keep mcont_pnais no matter what
      mutate(Diag = 1)
    don_reg_neg <- data_grp %>%
      uncount(non_pos) %>%
      select(a.inf, q.del, mcont_pnais, all_of(vars_reg)) %>%
      mutate(Diag = 0)
    don_reg <- don_reg_pos %>%
      bind_rows(don_reg_neg) %>%
      mutate(
        q.del = as.factor(q.del)) # default will put in numeric order
    
    # applying regression
   
    m <- glm(reg_form,
             data = don_reg,
             family = binomial(link = "cloglog")
    )
    
    ########## To get baseline distribution F for all covariates = 0
    
    time_coeffs <- m$coefficients[grepl("q.del", names(m$coefficients))] # these are alpha_j
    # this is 1-pj
    rev_p <- exp(-exp(time_coeffs))
    # to get F for covariates = 0, need cumulative product
    p_diag_cum = purrr::accumulate(rev(rev_p),
                                   .f = ~ .y * .x,
                                   .init = 1)
    p_diag_cum = rev(p_diag_cum)
    
    # store baseline F values
    F_mat <- bind_cols(F_baseline = p_diag_cum,
                       q.del = seq(0, length(p_diag_cum) - 1))
    
    ##### Work on variance of F_k
    # (For convenience, doing this before getting values of F_k)
    
    # Create a dataset, tmp, with all possible values of regression variables
    # q.del from 0 to m (kmax), and k_ind from 1 to m+2 (corresponding to a_1 to a_(m+2))
    # first format for q.del=0...
    data_grp_zeros <- data_grp_full %>%
      group_by(pick(all_of(vars_reg))) %>%
      summarise(q.del = 0) %>%
      ungroup() %>%
      mutate(k_ind=row_number())
    # ...then repeat and add q.del through max value m (kmax)
    tmp <- replicate(length(p_diag_cum) , data_grp_zeros, simplify = FALSE) %>%
      imap_dfr(~ .x %>% 
                 mutate(q.del = .y-1))
    # q.del=0 is not included in regression model because pjk=1
    tmp <- tmp %>%
      filter(q.del>0) %>% # can't have 0 here
      mutate(q.del=as.factor(q.del))
    # this gives values of a_1 through a_m ("a_val") for t=0
    # a_m is index of the output vector
    tmp$a_val <- predict(m, newdata=tmp, type="link")
    tmp <- tmp %>%
      mutate(a_val = -exp(a_val),
             a_ind = as.numeric(as.character(q.del))) %>%
      select(-c(q.del))
    # repeat again and add 'real' q.del (i.e. t=1, 2, ..., kmax)
    tmp <- replicate(length(p_diag_cum+1) , tmp, simplify = FALSE) %>%
      imap_dfr(~ .x %>% 
                 mutate(q.del = .y-1))
    # add zeros for q.del>0 
    tmp <- tmp %>%
      mutate(a_val = if_else(q.del>=a_ind , 0, a_val))
    # adding for betas
    betas <- tmp %>% group_by(q.del,pick(all_of(vars_reg)),k_ind) %>%
      summarise(tot = sum(a_val)) 
    # for factors, need this
    betas_mat <- as.data.frame(model.matrix(mm_form, betas)[,-1])
    beta_vars <- colnames(betas_mat)
    betas_mat <- betas_mat %>%
      select(-c(intersect(colnames(betas),colnames(betas_mat))))
    betas <- betas %>% bind_cols(betas_mat)
    # number of outputs corresponds to number of betas
    betas <- betas %>%
      pivot_longer(cols=all_of(beta_vars),
                   names_to="variable",
                   names_transform = list(variable = forcats::fct_inorder),
                   values_to="x_val") %>%
      mutate(a_ind = as.numeric(variable)+max(tmp$a_ind), # from m to m+(number of beta variales)
             a_val = x_val*tot) %>% # value = x*sum(a_1 to a_m)
      select(k_ind, a_ind, a_val, q.del) %>%
      full_join(data_grp_zeros %>% select(-c(q.del)))
    # size of vec_mat = (number of coefficients alpha and beta in glm model)*
    # (m+1 all possible delays)*(levels of X_k all possible covariates)
    vec_mat <- tmp %>%
      bind_rows(betas)
    
    # size = (number of coefficients alpha and beta in glm model)^2
    vcov_mat <- vcov(m)
    
    # To store variance of delay distributions
    F_var_mat <- vec_mat %>%
      group_by(q.del, k_ind,pick(all_of(vars_reg))) %>%
      arrange(a_ind) %>%
      summarise(var_log_F = as.numeric(a_val %*% vcov_mat %*% a_val)) %>%
      full_join(F_mat)
    
    
    ###### Dataset for incidence - rows per infection time, per imp
    
    don_merge_full <- data_strat %>%
      group_by(a.inf=a.inf.full,q.inf=q.inf.full,mcont_pnais) %>%
      summarise(n_diag_true = n()) %>%
      filter(a.inf>=min_year_diag)
    
    don_merge <- data_strat %>%
      group_by(a.inf, q.inf, q.diag, mcont_pnais) %>%
      summarise(n_diag = n()) %>%
      mutate(q.del = (month(max_quarter) - month(q.inf)) / 3 + (year(max_quarter) -
                                                                  year(q.inf)) * 4) %>%
      group_by(a.inf, q.inf, mcont_pnais) %>%
      summarise(n_diag = sum(n_diag), max_del = max(q.del)) %>%
      mutate(max_del = pmin(max_del, kmax)) %>%
      ungroup() %>%
      # un-truncated numbers of diagnoses for estimating 
      # size of the undiagnosed population
      left_join(don_merge_full) %>%
      mutate(n_diag_true = if_else(is.na(n_diag_true),0,n_diag_true)) %>%
      # years for regression
      mutate(
        year_2020 = if_else(a.inf == 2020, 1, 0),
        year_G20 = if_else(a.inf >= 2020, 1, 0),
        year_cont = a.inf - median(seq(min_year_diag,maxAn)),
        year_cont_L20 = if_else(a.inf < 2020, year_cont, 0),
        year_cont_G20 = if_else(a.inf >= 2020, year_cont, 0)
      ) 
    
    F_merged <- don_merge %>%
      left_join(F_var_mat %>% rename(max_del=q.del))  %>%
      ungroup()
    
    # Model coeffs that are not delays and not intercept
    model_coeffs <-
      m$coefficients[!grepl("q.del|Intercept*", names(m$coefficients))]
    print(model_coeffs)
    
    # Store coefficients for later use, if desired
    coeff_tmp <- bind_cols(names(model_coeffs),model_coeffs,rep(i,length(model_coeffs)))
    coeffs <- bind_rows(coeffs,coeff_tmp)
    
    # Applying coefficients to incidence matrix
    # To get F_k
    coeff_sum <- model.matrix(mm_form, F_merged)[,-1] %*% model_coeffs
    colnames(coeff_sum) <- 'coeff_sum'
    F_merged <- bind_cols(F_merged, coeff_sum) %>%
      mutate(F_final = F_baseline ^ exp(coeff_sum),
             var_F = F_final^2*var_log_F) 
    F_merged_new <- F_merged %>%
      mutate(
        Source = 'Brookmeyer_new',
        Inc = n_diag / F_final,
        Var_Inc = Inc * (1 - F_final) / F_final + Inc ^ 2 * var_F / F_final ^
          2,
        # Number still undiagnosed at the end of the period being considered
        Undiag = Inc - n_diag_true, # original approach
        Var_Undiag = Var_Inc # Inc - number diag (assumed known)
      ) %>%
      group_by(a.inf, Source) %>%
      summarise(Undiag = sum(Undiag), # FOR NUM DIAG
                Var_Undiag = sum(Var_Undiag),
                Inc = sum(Inc),
                Var_Inc = sum(Var_Inc)) %>% #assumed 0 covariance
      mutate(Inc_low = Inc - 1.96 * sqrt(Var_Inc),
             Inc_high = Inc + 1.96 * sqrt(Var_Inc),
             .imp=i, mcont_all=j)
    
    # in case of zeros, need to add back
    zeros <- expand.grid(unique(data_imp$a.inf),
                                unique(data_strat$mcont_all))
    colnames(zeros) <- c('a.inf','mcont_all')
    zeros <- zeros %>%
      mutate(Source='Brookmeyer_new', .imp=i)
    F_merged_new <- F_merged_new %>% full_join(zeros)
    F_merged_new <- F_merged_new %>% replace_na(list(Undiag = 0, Var_Undiag = 0,
                                                     Inc = 0, Var_Inc = 0))
    
    don_inc_est <- bind_rows(don_inc_est,F_merged_new)
  }
  
}

if (AccountForMig==T) {
  saveRDS(don_inc_est, paste0(PATH,'/don_inc_est.Rds'))
  saveRDS(pred_old, paste0(PATH,'/old_inf_inc.Rds'))
} else {
  saveRDS(don_inc_est, paste0(PATH,'/don_inc_est_nomig.Rds'))
  saveRDS(pred_old, paste0(PATH,'/old_inf_inc_nomig.Rds'))
}

########### Summarizing output

if (AccountForMig==T) {
  don_inc_est <- readRDS(paste0(PATH,'don_inc_est.Rds'))
  pred_old <- readRDS(paste0(PATH,'old_inf_inc.Rds'))
} else {
  don_inc_est <- readRDS(paste0(PATH,'don_inc_est_nomig.Rds'))
  pred_old <- readRDS(paste0(PATH,'old_inf_inc_nomig.Rds'))
}


# summarize across multiple imputations
don_inc_sum <- don_inc_est %>%
  group_by(annee.infection=a.inf, mcont_all) %>%
  summarise(Tot_Inc = pool.scalar(Q=Inc,U=Var_Inc)$qbar,
            Tot_Var = pool.scalar(Q=Inc,U=Var_Inc)$t,
            Tot_Undiag = pool.scalar(Q=Undiag,U=Var_Undiag)$qbar,
            Tot_Undiag_Var = pool.scalar(Q=Undiag,U=Var_Undiag)$t) %>%
  mutate(Inc_low = Tot_Inc - 1.96*sqrt(Tot_Var),
         Inc_high = Tot_Inc + 1.96*sqrt(Tot_Var)) %>%
  mutate(Inc = Tot_Inc,Source = 'Estimated')

# Undiagnosed - group all pre-min yr diag
undiag <- don_inc_sum %>%
  group_by(annee.infection,mcont_all) %>%
  summarise(Undiag = sum(Tot_Undiag),
            Undiag_Var = sum(Tot_Undiag_Var))
pred_old_merge <- pred_old %>%
  group_by(mcont_all) %>%
  summarise(Undiag = pool.scalar(Q=mean_diag,U=var_diag)$qbar,
            Undiag_Var = pool.scalar(Q=mean_diag,U=var_diag)$t) %>%
  mutate(annee.infection = min_year_diag-1)
undiag <- undiag %>%
  bind_rows(pred_old_merge) %>%
  mutate(Source='Estimated',
         Annee=2022) %>%
  group_by(Source,Annee,mcont_all) %>%
  summarise(Undiag=sum(Undiag),
            Undiag_Var = sum(Undiag_Var), 
            Undiag_Low = Undiag - 1.96*sqrt(Undiag_Var),
            Undiag_Hi = Undiag + 1.96*sqrt(Undiag_Var))

if (AccountForMig==T) {
  saveRDS(undiag, paste0(PATH,'/undiag_est_inc.Rds'))
} else {
  saveRDS(undiag, paste0(PATH,'/undiag_est_nomig.Rds'))
}

# Save incidence data in Excel
don_inc_excel <-  don_inc_sum %>%
  select(annee.infection, mcont_all,Inc, Inc_low, Inc_high) 
wb <- createWorkbook()
addWorksheet(wb,"Inc")
writeData(wb,sheet="Inc",don_inc_excel)
if (AccountForMig==T) {
  saveWorkbook(wb = wb, file = paste0(PATH,"inc_est_truemig.xlsx"), overwrite = T)  
} else {
  saveWorkbook(wb = wb, file = paste0(PATH,"inc_est_nomig.xlsx"), overwrite = T)  
  
}


#################################################################
#################################################################
################ Estimate HIV+ undiagnosed arrivals in France
################ And number undiagnosed, infected prior to arrival in France
#################################################################
#################################################################

# Technically don't need to redo this for no-migration versions
# but leaving in for simplicity because only takes about 2 min to run

don_diag_full <- readRDS(est_inf_dates)


########## Organizing the data 

don_diag_full <- don_diag_full %>%
  mutate(date.infection.est = date.diagnostic - dyears(DelayInfDiag)) %>%
  mutate(a.mig = annee.arriv, 
         a.diag = annee.diagnostic,
         a.del = a.diag - a.mig)  %>%
  mutate(mig.inf.est = if_else(date.arriv>date.infection.est,"PreMig","PostMig"),
         place.inf.est = if_else(pnais=="FRANCE"|mig.inf.est=="PostMig","FRANCE","OTHER")) %>%
  filter(place.inf.est != "FRANCE") %>%
  # place of birth and transmission route in 1 variable
  mutate(mcont_pnais = if_else(pnais=="FRANCE"&mcont=="HOMO-BI", "MSM_FRANCE", "MSM_ABROAD"),
         mcont_all = mcont_pnais)


min_diag=as.Date("2012-01-01"); 
max_diag=as.Date("2022-12-31");
min_year_diag=year(min_diag)
maxAn = year(max_diag) 


data <- don_diag_full %>%
  filter(annee.diagnostic<=maxAn)

kmax=10 # in years

don_inc_est <- NULL
coeffs <- NULL
pred_old <- NULL

set.seed(200)

n_imp <- max(data$.imp)
#n_imp <- 5 # TO TEST
for (i in 1:n_imp) {
  
  print(i)
  
  data_imp <- data %>% filter(.imp == i) 
  
  # Not adding "ALL" countries/regions of birth here
  # Because only 1 category for which relevant (MSM_abroad)
  # I suggest adding this if multiple categories
  #data_imp <- data_imp %>% 
  #  bind_rows(data_imp %>% mutate(mcont_all="ALL"))
  
  ##########################################################################
  #### Estimating diagnoses in people arrived prior to min_year_diag
  
  old_mig <- data_imp %>% filter(a.diag>=min_year_diag) %>% 
    group_by(a.diag,mcont_all) %>% 
    summarise(n_diag=n(), n_old=sum(a.mig<min_year_diag)) 
  
  # Going to impose that number migrated HIV+ pre-2012 diagnosed after 2022 
  # Can't exceed 3*number diagnosed during 2012-2022 (seems reasonable given 
  # median time to aids diagnosis ~11 years)
  curr <- old_mig %>%
    group_by(mcont_all) %>%
    summarise(max_old=3*sum(n_old))
  
  # roughly log-linear decline
  #ggplot(old_mig) +  geom_point(aes(x=a.diag,y=log(n_old)),col='red')+
  #  facet_wrap(vars(mcont_all),scales='free')
  
  # fit Poisson model
  # Don't include mcont_all because only 1 category (MSM_ABROAD); otherwise would
  m_old <- glm(n_old~a.diag,data=old_mig,family=poisson(link="log"))
  
  
  # transmission group categories, including "ALL"
  cat <- unique(data_imp$mcont_all)
  # dataset for predictions
  years <- sort(rep(seq(min_year_diag,min_year_diag+30),length(cat)))
  mcont <- rep(cat,length(seq(min_year_diag,min_year_diag+30)))
  pred_df <- bind_cols(a.diag=years,mcont_all=mcont)
  
  # To visually confirm how the model looks. 
  pred_df$n_old <- predict(m_old,pred_df,type="response")
  pred_df$Type <- 'Predicted'
  plot_df <- pred_df %>%
    bind_rows(old_mig %>% mutate(Type='Observed'))
  ggplot(plot_df) +
    geom_point(aes(x=a.diag,y=n_old,col=Type))+
    facet_wrap(vars(mcont_all),scales='free')
  ggplot(plot_df) +
    geom_point(aes(x=a.diag,y=log(n_old),col=Type))+
    facet_wrap(vars(mcont_all),scales='free') +
    ylab('Diagnoses of pre-2012 pre-migration infections') + 
    xlab('Year')
  #ggsave('old_mig_plot.png',height=4,width=7,units='in')
  
  
  # Bootstraps for within-imputation variance
  pred_df <- pred_df %>% 
    filter(a.diag>=year(max_diag))
  pred_boot <- Boot(m_old, f=function(x)predict(x, pred_df, type="response"), R=100,
                    method="case")
  pred_boot <- bind_cols(pred_df,t(pred_boot$t))
  pred_boot_sum <- pred_boot %>%
    group_by(mcont_all) %>%
    summarise(across(starts_with('...'), ~sum(.))) 
  pred_boot_sum$max_old <- curr$max_old # add max value for diag pre-2012
  pred_boot_sum <- pred_boot_sum %>%
    mutate(across(starts_with('...'), ~if_else(.>max_old,max_old,.))) 
  pred_boot_sum$mean_diag <- apply(pred_boot_sum %>% select(starts_with('...')),1,function(x) mean(x,na.rm=T))
  pred_boot_sum$var_diag <- apply(pred_boot_sum %>% select(starts_with('...')),1,function(x) var(x,na.rm=T))
  pred_boot_sum <- pred_boot_sum %>% select(mcont_all,mean_diag,var_diag) %>%
    mutate(.imp=i)
  pred_old <- bind_rows(pred_old,pred_boot_sum)
  
  ####################################################################
  ## Estimating arrivals and undiag for people arriving in [min_diag,max_diag]
  
  data_imp <- data_imp %>%
    mutate(a.del.full=a.del,
           a.mig.full=a.mig) %>%
    # Truncating for inc estimation only
    mutate(a.del = if_else(a.del>kmax, kmax, a.del),
           a.mig = a.diag - a.del) %>%
    mutate(a.trunc.del = year(max_diag) - a.mig) %>%
    filter(a.mig >= min_year_diag ) # needed for Brookmeyer
  
  # Start an inner loop here to estimate by route of transmission ('mcont')
  for (j in cat) {
    
    print(j)
    data_strat <- data_imp %>% filter(mcont_all==j)
    
    # Define regression formulas in advance
    
    # if all transmission groups, include variable mcont_pnais
    if (j=="ALL") { 
      vars_reg <- c('year_2020', 'year_cont', 'mcont_pnais') # Standard
      #vars_reg <- c('year_G20', 'year_cont_L20', 'mcont_pnais') # ECDC V1
      #vars_reg <- c('year_G20', 'year_cont_L20', 'year_cont_G20', 'mcont_pnais') # ECDC V2
      #vars_reg <- c('year_cont', 'mcont_pnais') # ECDC V3
    } else {
      vars_reg <- c('year_2020', 'year_cont') # Standard
      #vars_reg <- c('year_G20', 'year_cont_L20') # ECDC V1
      #vars_reg <- c('year_G20', 'year_cont_L20', 'year_cont_G20') # ECDC V2
      #vars_reg <- c('year_cont') # ECDC V3
    }
    
    
    reg_eqn <- paste(c("a.del",vars_reg),collapse="+") # for regression
    reg_form = as.formula(paste('Diag', "~0+", reg_eqn))
    
    mm_eqn <- paste(vars_reg,collapse="+") # for model matrix
    mm_form = as.formula(paste("~", mm_eqn))
    
    # if not at least 10 data points, with at least 2 different delays
    # Then error, insufficient data
    if (nrow(data_strat)<10 | length(unique(data_strat$a.del))<2) {
      stop("Too few data points to estimate migrations")
    }
    
    # Set first a.del = min(a.del) equal to zero
    # Will usually be true; if not, for small strata, may have issues dividing by zero
    data_strat$a.del[which(data_strat$a.del==min(data_strat$a.del))[1]] <- 0
    
    
    # Formatting for GLM                                               
    # First, creating grouped data
    # Stratifying variables will be mode of transmission/place of birth 
    # (mcont_pnais) and some form of year of migration (a.mig)
    
    # in case zeros
    mig_del_mat <- expand.grid(unique(data_imp$a.mig),
                               seq(0,kmax),unique(data_imp$mcont_pnais))
    colnames(mig_del_mat) <- c('a.mig','a.del','mcont_pnais') 
    
    data_grp <- data_strat %>%
      group_by(mcont_pnais, a.mig, a.del) %>%
      summarise(Yjk = n()) %>%
      full_join(mig_del_mat) %>%
      mutate(Yjk = replace_na(Yjk,0))
    
    # calculating njk (number at risk) by strata (a.mig) and delay
    # To calc nijk - first get number >=delay, then subtract those w/ trunc <delay
    # This is faster than doing in a loop
    num_Ldelay <- data_strat %>% # num w reporting delay <=a.del
      group_by(mcont_pnais,a.mig,a.del) %>% 
      summarise(n=n()) %>%
      full_join(mig_del_mat) %>%
      mutate(n = replace_na(n,0)) %>% 
      group_by(mcont_pnais,a.mig) %>%
      arrange(a.del) %>%
      mutate(n_a.del = cumsum(n)) 
    num_Ltrunc <- data_strat %>% # truncation time >= a.del
      group_by(mcont_pnais,a.mig,a.del=a.trunc.del) %>% 
      summarise(n=n()) %>%
      full_join(mig_del_mat) %>%
      mutate(n = replace_na(n,0)) %>% 
      group_by(mcont_pnais,a.mig) %>%
      arrange(a.del) %>%
      mutate(n_a.trunc = cumsum(n),
             n_a.trunc = n_a.trunc - n) %>% # for less than, not less than or equal
      select(-n) 
    num_all <- num_Ldelay %>%
      left_join(num_Ltrunc) %>%
      mutate(n_a.trunc=replace_na(n_a.trunc,0)) %>%
      mutate(njk=n_a.del-n_a.trunc) %>%
      select(a.mig,a.del,mcont_pnais,njk)
    data_grp <- data_grp %>%
      left_join(num_all)
    
    # estimating pjk and non_pos = njk-Yjk
    data_grp <- data_grp %>%
      mutate(
        year_2020 = if_else(a.mig == 2020, 1, 0),
        year_G20 = if_else(a.mig >= 2020, 1, 0),
        year_cont = a.mig - median(seq(min_year_diag,maxAn)),
        year_cont_L20 = if_else(a.mig < 2020, year_cont, 0),
        year_cont_G20 = if_else(a.mig >= 2020, year_cont, 0)
      )
    
    data_grp_full <- data_grp # incldues a.del=0, unlike below
    
    # estimating pjk and non_pos = njk-Yjk
    data_grp <- data_grp %>%
      mutate(pjk = Yjk / njk) %>%
      filter(njk > 0) %>%
      mutate(non_pos = njk - Yjk) %>% 
      filter(a.del != 0) %>% #because Yjk=njk
      ungroup() 
    
    # for regression, want 1 line per pos and 1 line per non-pos (per period at risk)
    don_reg_pos <- data_grp %>%
      uncount(Yjk) %>%
      select(a.mig, a.del, mcont_pnais, all_of(vars_reg)) %>%
      mutate(Diag = 1)
    don_reg_neg <- data_grp %>%
      uncount(non_pos) %>%
      select(a.mig, a.del, mcont_pnais, all_of(vars_reg)) %>%
      mutate(Diag = 0)
    don_reg <- don_reg_pos %>%
      bind_rows(don_reg_neg) %>%
      mutate(
        a.del = as.factor(a.del)) # default will put in numeric order
    
   
    # Applying regression
    m <- glm(reg_form,
             data = don_reg,
             family = binomial(link = "cloglog")
    )
    
    ########## To get baseline distribution F for coefficients = 0
    
    time_coeffs <- m$coefficients[grepl("a.del", names(m$coefficients))]
    # this is 1-pj
    rev_p <- exp(-exp(time_coeffs))
    # to get F for covariates = 0
    p_diag_cum = purrr::accumulate(rev(rev_p),
                                   .f = ~ .y * .x,
                                   .init = 1)
    p_diag_cum = rev(p_diag_cum)
    F_mat <- bind_cols(F_baseline = p_diag_cum,
                       a.del = seq(0, length(p_diag_cum) - 1))
    
    ##### Work on variance of F_k
    # (For convenience, doing this before getting values of F_k)
    
    # Create a dataset, tmp, with all possible values of regression variables
    # a.del from 0 to m (kmax), and k_ind from 1 to m+2 (corresponding to a_1 to a_(m+2))
    # (replacing 2 by the number of beta variables...)
    # first format for a.del=0...
    data_grp_zeros <- data_grp_full %>%
      group_by(pick(all_of(vars_reg))) %>%
      summarise(a.del = 0) %>%
      ungroup() %>%
      mutate(k_ind=row_number())
    # repeat and add a.del
    tmp <- replicate(length(p_diag_cum) , data_grp_zeros, simplify = FALSE) %>%
      imap_dfr(~ .x %>% 
                 mutate(a.del = .y-1))
    tmp <- tmp %>%
      filter(a.del>0) %>% # can't have 0 here (no a_0 in regression model)
      mutate(a.del=as.factor(a.del))
    # this gives values of a_1 through a_m
    tmp$a_val <- predict(m, newdata=tmp, type="link")
    tmp <- tmp %>%
      mutate(a_val = -exp(a_val),
             a_ind = as.numeric(as.character(a.del))) %>%
      select(-c(a.del))
    # repeat again and add real a.del
    tmp <- replicate(length(p_diag_cum+1) , tmp, simplify = FALSE) %>%
      imap_dfr(~ .x %>% 
                 mutate(a.del = .y-1))
    # add zeros for a.del>0 
    tmp <- tmp %>%
      mutate(a_val = if_else(a.del>=a_ind , 0, a_val))
    # adding for betas
    betas <- tmp %>% group_by(a.del,pick(all_of(vars_reg)),k_ind) %>%
      summarise(tot = sum(a_val)) 
    # for factors, need this
    betas_mat <- as.data.frame(model.matrix(mm_form, betas)[,-1])
    beta_vars <- colnames(betas_mat)
    betas_mat <- betas_mat %>%
      select(-c(intersect(colnames(betas),colnames(betas_mat))))
    betas <- betas %>% bind_cols(betas_mat)
    # number of outputs corresponds to number of betas
    betas <- betas %>%
      pivot_longer(cols=all_of(beta_vars),
                   names_to="variable",
                   names_transform = list(variable = forcats::fct_inorder),
                   values_to="x_val") %>%
      mutate(a_ind = as.numeric(variable)+max(tmp$a_ind),
             a_val = x_val*tot) %>%
      select(k_ind, a_ind, a_val, a.del) %>%
      full_join(data_grp_zeros %>% select(-c(a.del)))
    vec_mat <- tmp %>%
      bind_rows(betas)
    
    # GLM variance-covariance matrix
    vcov_mat <- vcov(m)
    
    # To store variance of delay distributions
    F_var_mat <- vec_mat %>%
      group_by(a.del, k_ind,pick(all_of(vars_reg))) %>%
      arrange(a_ind) %>%
      summarise(var_log_F = as.numeric(a_val %*% vcov_mat %*% a_val)) %>%
      full_join(F_mat)
    
    # Dataset for arrivals - rows per migration time, per imp
    don_merge_full <- data_strat %>%
      group_by(a.mig=a.mig.full,mcont_pnais) %>%
      summarise(n_diag_true = n()) %>%
      filter(a.mig>=min_year_diag)
    
    don_merge <- data_strat %>%
      group_by(a.mig, a.diag, mcont_pnais) %>%
      summarise(n_diag = n()) %>%
      mutate(a.del = year(max_diag) - a.mig) %>%
      group_by(a.mig, mcont_pnais) %>%
      summarise(n_diag = sum(n_diag), max_del = max(a.del)) %>%
      mutate(max_del = pmin(max_del, kmax)) %>%
      ungroup() %>%
      # un-truncated numbers of diagnoses for estimating number undiagnosed
      left_join(don_merge_full) %>%
      mutate(n_diag_true = if_else(is.na(n_diag_true),0,n_diag_true)) %>%
      # years for regression
      mutate(
        year_2020 = if_else(a.mig == 2020, 1, 0),
        year_G20 = if_else(a.mig >= 2020, 1, 0),
        year_cont = a.mig - median(seq(min_year_diag,maxAn)),
        year_cont_L20 = if_else(a.mig < 2020, year_cont, 0),
        year_cont_G20 = if_else(a.mig >= 2020, year_cont, 0)
      ) 
    
    F_merged <- don_merge %>%
      left_join(F_var_mat %>% rename(max_del = a.del))  %>%
      ungroup()
    
    # Model coeffs that are not delays and not intercept
    model_coeffs <-
      m$coefficients[!grepl("a.del|Intercept*", names(m$coefficients))]
    print(model_coeffs)
    
    # store coefficients for later, if desired
    coeff_tmp <- bind_cols(names(model_coeffs),model_coeffs,rep(i,length(model_coeffs)))
    coeffs <- bind_rows(coeffs,coeff_tmp)
    
    
    # Applying coefficients to incidence matrix
    coeff_sum <- model.matrix(mm_form, F_merged)[,-1] %*% 
      model_coeffs
    colnames(coeff_sum) <- 'coeff_sum'
    F_merged <- bind_cols(F_merged, coeff_sum) %>%
      mutate(F_final = F_baseline ^ exp(coeff_sum),
             var_F = F_final^2*var_log_F) 
    F_merged_new <- F_merged %>%
      mutate(
        Source = 'Brookmeyer_new',
        # Calling "inc", but truly, number of arriving immigrants with undiagnosed HIV
        Inc = n_diag / F_final,
        Var_Inc = Inc * (1 - F_final) / F_final + Inc ^ 2 * var_F / F_final ^
          2,
        # Number still undiagnosed at the end of the period being considered
        Undiag = Inc - n_diag_true, 
        Var_Undiag = Var_Inc #  Because subtracting a constant
      ) %>%
      group_by(a.mig, Source) %>%
      summarise(Undiag = sum(Undiag), # FOR NUM DIAG
                Var_Undiag = sum(Var_Undiag),
                Inc = sum(Inc),
                Var_Inc = sum(Var_Inc)) %>% #assumed 0 covariance
      mutate(Inc_low = Inc - 1.96 * sqrt(Var_Inc),
             Inc_high = Inc + 1.96 * sqrt(Var_Inc),
             .imp=i, mcont_all=j)
    
    
    # in case of zeros, need to add back
    zeros <- expand.grid(unique(data_imp$a.mig),
                         unique(data_strat$mcont_all))
    colnames(zeros) <- c('a.mig','mcont_all')
    zeros <- zeros %>%
      mutate(Source='Brookmeyer_new', .imp=i)
    F_merged_new <- F_merged_new %>% full_join(zeros)
    F_merged_new <- F_merged_new %>% replace_na(list(Undiag = 0, Var_Undiag = 0,
                                                     Inc = 0, Var_Inc = 0))
    
    
    don_inc_est <- bind_rows(don_inc_est,F_merged_new)
  }
  
}

saveRDS(don_inc_est,paste0(PATH,'don_mig_est.Rds'))
saveRDS(pred_old,paste0(PATH,'old_inf_mig.Rds'))

##### Summarizing for plots/tables

don_inc_est <- readRDS(paste0(PATH,'don_mig_est.Rds'))
pred_old <- readRDS(paste0(PATH,'old_inf_mig.Rds'))


# summarize across multiple imputations
# this is really arriving migrants
don_inc_sum <- don_inc_est %>%
  filter(mcont_all!="ALL") %>% # ALL = MSM_ABROAD here, so no need
  group_by(annee.migration=a.mig, mcont_all) %>%
  summarise(Tot_Arriv = pool.scalar(Q=Inc,U=Var_Inc)$qbar,
            Tot_Var = pool.scalar(Q=Inc,U=Var_Inc)$t,
            Tot_Undiag = pool.scalar(Q=Undiag,U=Var_Undiag)$qbar,
            Tot_Undiag_Var = pool.scalar(Q=Undiag,U=Var_Undiag)$t) %>%
  mutate(Arriv_low = Tot_Arriv - 1.96*sqrt(Tot_Var),
         Arriv_high = Tot_Arriv + 1.96*sqrt(Tot_Var)) %>%
  mutate(Arriv = Tot_Arriv,Source = 'Estimated')

# Undiagnosed - group all pre-min yr diag
undiag <- don_inc_sum %>%
  group_by(annee.migration,mcont_all) %>%
  summarise(Undiag = sum(Tot_Undiag),
            Undiag_Var = sum(Tot_Undiag_Var))
pred_old_merge <- pred_old %>%
  group_by(mcont_all) %>%
  filter(mcont_all!="ALL") %>% # ALL = MSM_ABROAD here, so no need
  summarise(Undiag = pool.scalar(Q=mean_diag,U=var_diag)$qbar,
            Undiag_Var = pool.scalar(Q=mean_diag,U=var_diag)$t) %>%
  mutate(annee.migration = min_year_diag-1)
undiag <- undiag %>%
  bind_rows(pred_old_merge) %>%
  mutate(Undiag_Low = Undiag - 1.96*sqrt(Undiag_Var), 
         Undiag_Hi = Undiag + 1.96*sqrt(Undiag_Var),
         Source='Estimated',
         Annee=2022) %>%
  group_by(Source,Annee,mcont_all) %>%
  summarise(Undiag=sum(Undiag),
            Undiag_Var = sum(Undiag_Var), 
            Undiag_Low = Undiag - 1.96*sqrt(Undiag_Var),
            Undiag_Hi = Undiag + 1.96*sqrt(Undiag_Var))

saveRDS(undiag, paste0(PATH,'/undiag_est_mig.Rds'))

# Save immigration estimates in Excel
don_inc_excel <-  don_inc_sum %>%
  select(annee.migration, mcont_all,Arriv, Arriv_low, Arriv_high) 
wb <- createWorkbook()
addWorksheet(wb,"Immig")
writeData(wb,sheet="Immig",don_inc_excel)
saveWorkbook(wb = wb, file = paste0(PATH,"mig_est.xlsx"), overwrite = T)  

#################################################################
#################################################################
################ Size of the undiagnosed population
################ Combining those infected in France and migrants
#################################################################
#################################################################

wb <- createWorkbook()
addWorksheet(wb,"Undiag")

if (AccountForMig==T) {
  undiag_inc <- readRDS(paste0(PATH,'/undiag_est_inc.Rds'))
  undiag_inc <- undiag_inc %>% mutate(Source='Inf')
  undiag_mig <- readRDS(paste0(PATH,'/undiag_est_mig.Rds'))
  undiag_mig <- undiag_mig %>%
    # because only 1 category, did not previous add 'ALL'
    bind_rows(undiag_mig %>% mutate(mcont_all="ALL")) %>%
    mutate(Source = 'Mig')
  undiag_all <- undiag_inc %>%
    bind_rows(undiag_mig) %>%
    group_by(Annee,mcont_all) %>%
    summarise(Undiag = sum(Undiag),
              Undiag_Var = sum(Undiag_Var), 
              Undiag_Low = Undiag - 1.96*sqrt(Undiag_Var),
              Undiag_Hi = Undiag + 1.96*sqrt(Undiag_Var))
  writeData(wb,sheet="Undiag",undiag_all)
  saveWorkbook(wb = wb, file = paste0(PATH,"undiag_est_truemig.xlsx"), overwrite = T)  
  
  
} else {
  undiag_all <- readRDS(paste0(PATH,'/undiag_est_nomig.Rds'))
  writeData(wb,sheet="Undiag",undiag_all)
  saveWorkbook(wb = wb, file = paste0(PATH,"undiag_est_nomig.xlsx"), overwrite = T)  
  
}
```
