## Supplemental Data and Programmes for "A new approach to estimating HIV incidence and the size of the infected and undiagnosed population in high immigration settings": Estimate_Infection_Dates_Final.R

```
# Part I: estimate infection times

# Adapted from codes in supplementary material of:
# Pantazis N, Thomadakis C, Del Amo J, Alvarez-Del Arco D, Burns FM, Fakoya I, Touloumi G. 
# Determining the likely place of HIV acquisition for migrants in Europe combining 
# subject-specific information and biomarkers data. Stat Methods Med Res. 
# 2019 Jul;28(7):1979-1997. doi: 10.1177/0962280217746437.

library(tidyverse)
library(lubridate)
library(mvtnorm)
library(splines)
library(purrr)
library(foreach)
library(mice)
library(doRNG) # reproducible parallel computation

####################################################################
####################################################################
##### Functions required 
##### Modified from Pantazis et al (2019)
####################################################################
####################################################################


### # w = delay, don_row = data, pars_all = a list of parameters for all rows
# data must have variables 
#"gender","region_group","expo","ageDiag","dtime","calendar",
# "consc","yvar", "u", "beta_hat" 
### Includes expression for competing risks: Data must include column beta_hat
#### Version using CD4 only, not VL
log_postWcd4_ak2 = function(w,don_row,pars_all)
{
  
  if (w<0.01-1e-6 | w > don_row$u+1e-6 ) {
    return(p=Inf) # negative log lik
  } else {
    
    don_row <- don_row %>% mutate(age_inf = ageDiag - w)
    # Ignoring mig with pre-migration odds etc
    
    faids = formula(id ~ gender + region_group + expo + age_inf)
    x.aids = model.matrix(faids,don_row)
    maxdtime = 0 # only observing at time of diagnosis
    kappa = pars_all$kappa
    lambda = exp(x.aids %*% pars_all$beta.aids)
    
    # Formula for design matrix of fixed effects
    fxcd4 = formula(yvar ~ I(dtime+w)*gender + I(dtime+w)*region_group + I(dtime+w)*expo 
                    + I(dtime+w)*lspline(I(ageDiag-w),knots = c(25,35,45)) 
                    + lspline(I(calendar-w),knots = c(16,22)))
    x = model.matrix(fxcd4,data = don_row)  
    
    # Formula for design matrix of random effects
    fz = formula(yvar ~ -1 + consc + I(dtime+w) )  
    z = model.matrix(fz,data = don_row)
    
    # Mean and variance of the normal kernel
    mu = c(x%*%pars_all$bFE.CD4)
    var = z%*%tcrossprod(pars_all$VarCovRE.CD4,z) + pars_all$sigma2.CD4*diag(length(x[,1]))
    
    p = dmvnorm(don_row$yvar,mean = mu, sigma = var,log = TRUE) - lambda*(w + maxdtime)^kappa
    
    p = p + (log(kappa) + log(lambda) + (kappa - 1)*log(w + maxdtime))*(don_row$sida) - (1/don_row$beta_hat)*w
    
    return(-p)
  }
}


postWcd4_ak2 = function(w,don_row,pars_all)
{
  return(exp(-log_postWcd4_ak2(w,don_row,pars_all)))
}


# Simulate random draws from the posterior distribution 
# with function posteriorW, mode pmode
# other variables from don_row and pars_all
rejection.sim_ak = function(n,posteriorW,pmode,don_row,pars_all)
{
  # Rejection sampling
  # n: number of accepted simulations
  # it: index for iterations
  # accept: index for accepted values
  
  it = 0
  accept = 0
  
  # Sample: vector with accepted values
  
  sample = rep(0,n)
  K = posteriorW(pmode,don_row,pars_all)
  K=as.vector(K) # K is 1x1 but get a warning if don't do this
  
  while (accept<n)
  {
    it = it+1
    
    # Envelope function g(theta)~U(0,1)
    u = runif(n - accept,don_row$low,don_row$u) 
    v = runif(n - accept)
    post = sapply(u,posteriorW,don_row,pars_all) 
    if ( any(v < post/K) )
    {
      a = v < post/K
      g = sum(a)
      sample[(accept+1):(accept+g)] = u[a]
      accept = accept + g
    } 
  }
  # Output list
  out = list()
  out$sample = sample
  out$it = c("Iterations required:",it)
  out$accept = c("Acceptance rate",n/it)
  
  return(out)
} 

####################################################################
### Function implementing linear splines                         ###
### Obtained from the "ares" package (old could not be used)     ###
### Very minor changes were made, mainly some ifs  were dropped  ###
####################################################################

lspline = function (var, knots = NULL, nknots = NULL, percentiles = NULL, 
                    marginal = FALSE, names = NULL) 
{
  n <- length(var)
  if (!is.null(nknots)) 
    if ((nknots > n/2) || (length(knots) > n/2) || (length(percentiles) > 
                                                    n/2)) 
      stop("Less than two points between two knots")
  if (is.null(nknots) && is.null(knots) && is.null(percentiles)) 
    stop("Either knots position, number of knots, or percentiles for the knots must be supplied")
  if (sum(!is.null(nknots), !is.null(knots), !is.null(percentiles)) > 
      1) 
    stop("Knots position, number of knots and percentiles are mutually excludent")
  if (!is.null(knots)) {
    
    w <- "knots"
    k <- length(knots) + 1
  }
  else if (!is.null(percentiles)) {
    w <- "percentiles"
    k <- length(percentiles) + 1
  }
  else {
    w <- "nknots"
    k <- nknots + 1
  }
  if (w == "nknots") 
    knots <- quantile(var, probs = seq(0, 1, by = 1/k)[2:k], 
                      na.rm = TRUE)
  else if (w == "percentiles") 
    knots <- quantile(var, probs = percentiles, na.rm = TRUE)
  knots <- c(knots, max(var, na.rm = TRUE))
  if (marginal) {
    new.vars <- var
    for (i in 2:k) {
      tmp.var <- double(n)
      tmp.var <- pmax(0, (var - knots[i - 1]))
      new.vars <- cbind(new.vars, tmp.var)
    }
  }
  else {
    new.vars <- pmin(var, knots[1], na.rm = FALSE)
    for (i in 2:k) {
      tmp.var <- double(n)
      tmp.var <- pmax(pmin(var, knots[i], na.rm = FALSE), 
                      knots[i - 1], na.rm = FALSE) - knots[i - 1]
      new.vars <- cbind(new.vars, tmp.var)
    }
  }
  if (length(names) == k) 
    dimnames(new.vars)[[2]] <- names
  else dimnames(new.vars)[[2]] <- paste(deparse(substitute(var)), 
                                        ".", 1:k, sep = "")
  retval <- new.vars
  class(retval) <- "lspline"
  return(retval)
}


############################################################
############################################################
######### Estimating infection dates
############################################################
############################################################


###############################################
### Get the estimates from the CASCADE data ###
### Will need to edit names/directories/etc as needed
###############################################

WR = './Params/'

# Estimates of the fixed effects parameters from the bivariate LMM applied to CASCADE data
FixedEffectsBivCASCADE = read.csv(paste0(WR,"FixedEffectsBivCASCADE_pant.csv"))
bFE = FixedEffectsBivCASCADE[,1]
names(bFE) = rownames(FixedEffectsBivCASCADE)

names(bFE)[1:27]
bFE.CD4 = bFE[1:27]

names(bFE)[28:length(bFE)]
bFE.VL = bFE[28:length(bFE)]

# and variance-covariance parameters
VarCovBivCASCADE = read.csv(paste0(WR,"VarCovBivCASCADE_pant.csv"))
VarCovRE = as.matrix(VarCovBivCASCADE[1:5,1:5])
sigma2 = unlist(c(VarCovBivCASCADE[1,6:7]))
sigma2.CD4 = sigma2[1]
sigma2.VL = sigma2[2]
VarCovRE.CD4 = VarCovRE[1:2,1:2]
VarCovRE.VR = VarCovRE[3:5,3:5]


# Parameters regarding the time-to-AIDS model to be used
estTimeToAIDS = read.csv(paste0(WR,"estTimeToAIDS_pant.csv"))
beta.aids = estTimeToAIDS[-nrow(estTimeToAIDS),1]
names(beta.aids) = rownames(estTimeToAIDS)[-nrow(estTimeToAIDS)]
kappa = estTimeToAIDS[nrow(estTimeToAIDS),1]
names(kappa) = "kappa"

#################################
########## Upload (simulated) data
######### Run separately for COVID and non-COVID data
##################################

COVID_drop_diag = F # T F

if (COVID_drop_diag==T) {
  don_samp <- readRDS('bdd_testeurs_covid_mig.Rds')
} else {
  don_samp <- readRDS('bdd_testeurs_mig.Rds')
}


############################
## Format data for model
############################

don_diag_wide= don_samp %>%
  # Assume we have surveillance data from 2012-2022
  filter(annee.diagnostic >= 2012 & annee.diagnostic < 2023) %>%
  # Defining longest possible infection
  # Simulated data starts at 16yo, so set that as min age
  mutate(u = pmin(age-16+0.5, # Assign half a year of risk to age 16
                  annee.diagnostic-1980+0.5)) %>% # Similar to 1980
  mutate(low = 0.01) %>%
  mutate(u = if_else(testeur=="rep", pmin(u,(delnegpos_m+0.5)/12),u)) # Delays from last neg to first pos test are in months

# Check for non-sim data; should be zero
don_diag_wide %>% filter(u<=low) %>% nrow()

# For "fast" version, round variables
don_diag_wide <- don_diag_wide %>%
  mutate(u_cat = if_else(u>=5,round(u), # to nearest year if >5 years
                         ceiling(u*4)/4))  %>% # to nearest quarter (rounding up) if <5 years
  mutate(u = u_cat) %>%
  mutate(
    cd4_50 = round(nbt4_m*2,-2)/2, # to nearest 50, max 1500
    cd4_50 = if_else(cd4_50>=1500,1500,cd4_50),
    gender = sexe,
    region_group = if_else(pnais=="FRANCE","EUROPE",pnais),
    expo = mcont,
    sida = sida ,
    age_5 = round(age*2,-1)/2, # to nearest 5, max 80
    age_5 = if_else(age>=80,80,age_5),
    calendar_5 = round((annee.diagnostic-1980)*2,-1)/2 #to nearest 5
  )

######### Add rows to database
######### As if multiple imputations
don_diag_wide <- replicate(15, don_diag_wide, simplify = FALSE) %>%
  imap_dfr(~ .x %>%
             mutate(.imp = .y))

# Adding params for competing risks.
# Real data, do by mcont, so keep that structure here
crisks <- don_diag_wide %>%
  filter(testeur=="primo") %>% #first-time testers
  #filter(stclin_m!="PIV") %>% #not in primary infection
  group_by(mcont) %>%
  summarise(n=n(),
            n_sida=sum(stclin_m=="SID")) %>%
  mutate(p_sida=n_sida/n) # proportion diagnosed at AIDS stage
alpha_a = 5.251; beta_a = 1.974
crisks <- crisks %>%
  mutate(beta_hat = beta_a/(p_sida^(-1/alpha_a)-1))
# do a left_join if include mcont later
don_diag_wide <- don_diag_wide %>%
  left_join(crisks %>% select(mcont, beta_hat))
# assume primary infection and repeat testers follow different dynamics, so set beta_hat
# high enough to have no impact
don_diag_wide <- don_diag_wide %>%
  mutate(beta_hat = if_else(testeur=="rep"|stclin_m=="PIV",100000,beta_hat))


# Grouped data.
# Speed gains greater for real database than the smaller simulated one
don_diag_wide_indiv <- don_diag_wide
don_diag_wide = don_diag_wide %>%
  group_by(cd4_50,gender,region_group,expo,age_5,
           sida,u=u_cat,calendar_5, beta_hat, low) %>%
  summarise(n_diag_cat=n()) %>%
  ungroup() %>%
  mutate(cat_num = row_number())

## Adding variables
don_diag_wide = don_diag_wide %>%
  mutate(id = row_number(),
         ageDiag = age_5,
         calendar = calendar_5)

# Declare variables as factors. Must be done to match the corresponding estimates for the population parameters
don_diag_wide$gender = factor(don_diag_wide$gender,levels = c("F","M")) # order must be Female, Male
don_diag_wide$region_group = factor(don_diag_wide$region_group,levels = c("AFSS","EUROPE","ASIA","AUTRE")) # order must be Africa, Europe, Asia, America/Other
don_diag_wide$expo = factor(don_diag_wide$expo,levels = c("HOMO-BI","UDI","HETERO","AUTRE")) # order must be MSM, IDU, Hetero, Other

######## Variables for CD4 only
don_diag_cd4 = don_diag_wide %>%
  mutate(yvar = cd4_50^(1/4), # Fourth root CD4, version Pantazis 2019
         consc=1, consr=0,
         dtime=0 # time between diagnosis and VL/CD4
  )
don_diag_wide <- don_diag_cd4


# returning to individual level format, also need to redo factors
don_diag_wide_grp <- don_diag_wide
don_diag_wide <- don_diag_wide_indiv %>%
  left_join(don_diag_wide)
don_diag_wide$gender = factor(don_diag_wide$gender,levels = c("F","M")) # order must be Female, Male
don_diag_wide$region_group = factor(don_diag_wide$region_group,levels = c("AFSS","EUROPE","ASIA","AUTRE")) # order must be Africa, Europe, Asia, America/Other
don_diag_wide$expo = factor(don_diag_wide$expo,levels = c("HOMO-BI","UDI","HETERO","AUTRE")) # order must be MSM, IDU, Hetero, Other

### Correspondence between grouped and individual datasets
don_diag_wide <- don_diag_wide %>%
  arrange(cat_num)
# number of people in each group
draw_mat <- don_diag_wide %>%
  mutate(ind=row_number()) %>%
  group_by(cat_num,n_diag_cat) %>%
  summarise(min_row=min(ind),max_row=max(ind))

print(nrow(don_diag_wide))

############################################
##### Estimating infection times - parallelized
##### This version takes about 18minutes
###########################################

set.seed(100) # for reproducibility

time1<-Sys.time()
pars_all <- list(beta.aids=beta.aids,bFE.CD4=bFE.CD4,
                 VarCovRE.CD4=VarCovRE.CD4,sigma2.CD4=sigma2.CD4,
                 kappa=kappa)
cl <- parallel::makeCluster(4)
doParallel::registerDoParallel(cl)
draws <- foreach(i = 1:nrow(don_diag_wide_grp),
                 .combine="rbind",
                 .packages=c('tidyverse','lubridate', 'mvtnorm','splines','purrr'),
                 .export=c('lspline')) %dorng% {
                   # using optimize instead of optim removes risk of convergence issue
                   fit = optimize(log_postWcd4_ak2,
                                  interval = c(don_diag_wide_grp$low[i],don_diag_wide_grp$u[i]),
                                  don_row=don_diag_wide_grp[i,],pars_all=pars_all)
                   ss = rejection.sim_ak(draw_mat$n_diag_cat[i],pmode = fit$minimum,
                                         posteriorW=postWcd4_ak2,
                                         don_row=don_diag_wide_grp[i,],pars_all=pars_all)
                   output = bind_cols(rep(i,length(ss$sample)),
                                      ss$sample)
                   return(output)
                 }
colnames(draws) <- c("id2","DelayInfDiag")
parallel::stopCluster(cl)
time2<-Sys.time()
time2-time1
don_diag_wide <- don_diag_wide %>%
  bind_cols(draws)
# should be no rows here if works correctly
don_diag_wide %>% filter(id!=id2) %>% nrow()

don_diag_full = don_diag_wide %>% select(-c(id2))


if (COVID_drop_diag==T) {
  saveRDS(don_diag_full,"sim_output_fast_covid.Rds")
} else {
  saveRDS(don_diag_full,"sim_output_fast_nocovid.Rds")
}
```
