## Supplemental Data and Programmes for "A new approach to estimating HIV incidence and the size of the infected and undiagnosed population in high immigration settings": export_sim_ecdc.R

```
# Export simulated data for ECDC model
# Assuming place of infection always = France (i.e. no pre-migration infections)

library(tidyverse)

# COVID version
don_samp <- readRDS('bdd_testeurs_covid_mig.Rds')
PATH = './ECDC_export_covid/'

# No COVID version
# don_samp <- readRDS('bdd_testeurs_mig.Rds')
# PATH = './ECDC_export_nocovid/'

don_samp <- don_samp %>%
  mutate(year=annee.diagnostic)

export1_inc_hiv <- don_samp %>%
  filter(year>=2003 & year<=2022) %>%
  group_by(year) %>%
  summarise(ALL=n()) 
write.csv(export1_inc_hiv,paste0(PATH,'HIV.csv'),row.names=F) 


export1_inc_hivaids <- don_samp %>%
  filter(year>=2003 & year<=2022 & sida==1) %>%
  group_by(year) %>%
  summarise(ALL=n()) 
write.csv(export1_inc_hivaids,paste0(PATH,'HIVAIDS.csv'),row.names=F) 

export1_inc_cd41 <- don_samp %>%
  filter(year>=2008 & year<=2022 & sida==0 & nbt4_m>=500) %>%
  group_by(year) %>%
  summarise(ALL=n()) 
write.csv(export1_inc_cd41,paste0(PATH,'HIV_CD4_1.csv'),row.names=F) 

export1_inc_cd42 <- don_samp %>%
  filter(year>=2008 & year<=2022 & sida==0 & nbt4_m>=350 & nbt4_m<500) %>%
  group_by(year) %>%
  summarise(ALL=n()) 
write.csv(export1_inc_cd42,paste0(PATH,'HIV_CD4_2.csv'),row.names=F) 

export1_inc_cd43 <- don_samp %>%
  filter(year>=2008 & year<=2022 & sida==0 & nbt4_m>=200 & nbt4_m<350) %>%
  group_by(year) %>%
  summarise(ALL=n()) 
write.csv(export1_inc_cd43,paste0(PATH,'HIV_CD4_3.csv'),row.names=F) 

export1_inc_cd44 <- don_samp %>%
  filter(year>=2008 & year<=2022 & sida==0 & nbt4_m<200) %>%
  group_by(year) %>%
  summarise(ALL=n()) 
write.csv(export1_inc_cd44,paste0(PATH,'HIV_CD4_4.csv'),row.names=F) 

# Just forcing everyone to 0 in 1980 
export1_inc_aids <- don_samp %>% 
  mutate(year=1980) %>%
  group_by(year) %>%
  summarise(ALL=0)
write.csv(export1_inc_aids,paste0(PATH,'AIDS.csv'),row.names=F)

#######################################################
##### Step 2: Run HIV modelling tool online
##### https://shinyapps.ecdc.europa.eu/shiny/hivPlatform/
##### Access date 13 Sept 2024 (Version 3.0.3, Build Date: 2024-06-22)
##### Upload aggregated data
#####
##### Options - 
# Data since start of pandemic: no
# Knots: 6
# Start at zero: yes
# Prevent sudden changes at end: yes
# ML distribution: Poisson
# Output "detailed main fit model results" and "detailed bootstrap fits model results" 
# Migrant connection: NO
# 100 parametric bootstrap iterations
#
# Version 1 Diagnosis matrix:
# - 1980-1984: no boxes checked
# - 1984-2003: jump only
# - 2003-2010: jump and change by CD4 count
# - 2010-2020: change by CD4 count and change in interval
# - 2020-2023: jump and change by CD4 count
#
# Version 2 Diagnosis matrix:
# - 1980-1984: no boxes checked
# - 1984-2003: jump only
# - 2003-2010: jump and change by CD4 count
# - 2010-2020: change by CD4 count and change in interval
# - 2020-2023: jump and change by CD4 count and change in interval
#
# Version 3 Diagnosis matrix:
# - 1980-1984: no boxes checked
# - 1984-2003: jump only
# - 2003-2012: jump and change by CD4 count
# - 2012-2023:  change by CD4 count and change in interval
```
