## Supplemental Data and Programmes for "A new approach to estimating HIV incidence and the size of the infected and undiagnosed population in high immigration settings": README.docx

The contents of this folder are as follows.

Empty folders, used to store outputs:

- Covid
- Nocovid
- ECDC_export_covid
- ECDC_export_nocovid

Datasets:

- **bdd_testeurs_covid_mig.Rds** and **bdd_testeurs_mig.Rds**: these contain the two datasets described in the article, one that assumes a temporary drop in the probability of diagnosis in 2020-2021 (“bdd_testeurs_covid_mig.Rds”) and the other without this COVID-associated drop in diagnoses (“bdd_testeurs_mig.Rds”). To test the data on simulate data “without migration”, we used the same datasets but ignoring all variables related to migration, and thus assuming all people were infected within the country of interest. The variables of these datasets are as follows:
  - date.infection: Date of infection
  - annee.infection: Year of infection
  - date.diagnostic: Year of HIV diagnosis
  - annee.diagnostic: Year of HIV diagnosis
  - age: Age in years at the time of diagnosis
  - log_cv_m: Measured viral load, log10, at the time of diagnosis
  - nbt4_m: Measured CD4 count at the time of diagnosis
  - sida: Whether the person has AIDS or not at the time of diagnosis
  - cdcb: Whether the person has CDC Category B symptoms at the time of diagnosis
  - tb: Whether the person has tuberculosis at the time of diagnosis
  - stclin_m: Clinical stage at the time of diagnosis. Options are ASY (asymptomatic), PIV (primary HIV infection), SID (AIDS), or SNS (symptomatic non-AIDS). A value of 0 is assigned to people diagnosed after 2022.
  - testeur: Whether the person ever had a previous negative HIV test (“rep”) or not (“primo”) prior to HIV diagnosis
  - serneg: Late of last negative HIV test, if applicable
  - delnegpos_m: Months elapsed between the person’s last negative HIV test and first positive HIV test
  - cd4_cat: CD4 at the time of diagnosis, in categories
  - delinfpos_j: days between infection and first positive test
  - pnais: place of birth (options: “FRANCE” or “EUROPE” if outside France)
  - sexe: M (always male)
  - mcont: Route of transmission. Always HOMO-BI (sexual transmission between MSM)
  - date.arriv: Date of immigration
  - annee.arriv: Year of immigration
- **Folder “Params”**: these files contain the parameters necessary to estimate infection times. They come from the supplementary appendix to Pantazis N, Thomadakis C, Del Amo J, Alvarez-Del Arco D, Burns FM, Fakoya I, Touloumi G. Determining the likely place of HIV acquisition for migrants in Europe combining subject-specific information and biomarkers data. Stat Methods Med Res. 2019 Jul;28(7):1979-1997. doi: 10.1177/0962280217746437.

Codes:

- **export_sim_ecdc.R**: Code used to export the simulated data in the format required by the ECDC HIV Modeling Tool. Also contains the information about how this tool was accessed and the parameters used.
- **Estimate_Infection_Dates_Final.R:** Creates simulated datasets augmented with 15 (index “.imp”) estimated delays between infection and diagnosis per person (variable “DelayInfDiag”). You need to specify whether you want this to run on the dataset including a COVID-associated drop in diagnosis probabilities from 2020-2021 or not (variable “COVID_drop_diag” = T or F, line 230). The output is called “sim_output_fast_covid” or "sim_output_fast_nocovid.Rds". In our experience this code takes approximately 20 minutes to run on each simulate dataset. Note: This code must be run before Estimate_Incidence_Undiag_Final.R.
- **Estimate_Incidence_Undiag_Final.R**: Reads in the output from Estimate_Infection_Dates_Final.R. You need to specify whether estimates should be made accounting for migration (“AccountForMig = T”) or ignoring migration (“AccountForMig = F”), and which dataset to use. The default includes the “original” model assuming a diagnosis delays change linearly from 2012 to 2022 with a temporary drop in 2020. Parameters needed for the other tested parameter sets are also included. In our experience this code takes around 10 minutes to run on each simulated dataset per parameter set. Outputs save to the folders “Covid’ or “Nocovid”, according to the simulated dataset used, and include:
  - True simulated values of the size of the undiagnosed population in 2022 (True_Sim_Undiag.xlsx), and HIV incidence and the number of immigrants arriving with undiagnosed HIV from 2012-2022 (True_Sim_Inc_Mig.xlsx)
  - New model estimates of HIV incidence (“inc_est_...xlsx”), immigration of people with undiagnosed HIV (“mig_est.xlsx”), and the number of people with undiagnosed HIV infection at the end of 2022 (“undiag_est…xlsx”) as well as intermediate files.
